# Effects of statin treatment on primary and hospital care use: a microsimulation model

**DOI:** 10.1101/2025.09.30.25337016

**Authors:** Junwen Zhou, Runguo Wu, Claire Williams, Chris Carvalho, John Robson, Borislava Mihaylova

## Abstract

**Background:** Statin treatment’s efficacy, safety and cost-effectiveness are well accepted, but the long-term impact on healthcare use is not fully understood. We assessed statin treatment effects on primary and hospital care use over time.

**Methods:** The UK Biobank population cohort with linked hospital admissions (N=501,807) and primary healthcare data (N=192,983) informed models of hospital admissions, hospital inpatient days, primary care services (consultations, diagnostic and monitoring tests, and medication prescription items) associated with individual characteristics and occurrences of myocardial infarction, stroke, coronary revascularization and vascular death. These models were integrated into a validated cardiovascular disease (CVD) microsimulation policy model to assess statin treatment effects on healthcare use in population categories by age (40-60 years and 60-70 years) and prior CVD history.

**Results:** Statin treatment was associated with improved survival and lower rates of hospital admissions, hospital inpatient days and prescription items per person-year over lifetime. Compared to no treatment, healthcare use with statin treatment was lower in earlier years after initiation, these reductions diminished over time and transitioned into higher healthcare use. The number of years to net neutral effect (95%CI) ranged from 9 (7-12) to 17 (9-25) for consultations/tests, from 22 (17-28) to 38 (28-48) for prescription items/hospital admissions, and from 40 (30-47) to 51 (43-62) for hospital inpatient days. Earlier transitions were observed in older people and people with prior CVD history.

**Conclusions:** Statin treatment reduces individual rates of hospital inpatient services and medication prescriptions but increases overall healthcare use driven by increased longevity and ageing.

**Funding:** NIHR-HTA, NIHR-Barts-BRC.

## Introduction

Cardiovascular diseases (CVDs) are the leading causes of death worldwide, contributing to substantial health and economic burden globally^1^. In the United Kingdom (UK), 7.6 million people are living with CVD, which is associated with 0.17 million deaths and £12 billion healthcare costs annually^2^. Statin therapy is widely recommended to reduce CVD events in people at increased risk^3-6^. Evidence on the efficacy^4^ and safety^7^ of statins has informed clinician-patient discussions of statin treatment^3^ and, alongside evidence on statins’ effects on quality-adjusted life years, healthcare costs and cost-effectiveness^5,6^, has supported widening guideline recommendations for statin use^3^.

However, impact of statin treatment on healthcare use has received less attention. This is important for both healthcare planning and resource allocation as well as for patients themselves. While preventive interventions lower the risk of CVD, they increase survival and may increase overall healthcare use due to healthcare need during extended life years and increased morbidity with ageing. The need for healthcare due to ill health also substantially affects patients’ lives consuming time, energy and financial resources and causing personal discomfort^8,9^.

In this study, we assessed the impact of CVD events on hospital admissions and primary care services in the UK and evaluated the impact of statin therapy on healthcare use using a CVD microsimulation model.

## Methods

### Study population and data

The UK Biobank (UKB) study is a prospective cohort study of over 500,000 adults, aged 40-70 years at recruitment from 2006 to 2010 across England, Scotland, and Wales^10^. All participants with established linkage to primary (40% of the cohort) or hospital care records, except a small number at end-stage kidney disease at recruitment, contributed to our current study (**Table S1**). Participants’ data from enrolment into UKB until the earliest of 31 March 2016, death or loss to follow-up, contributed to the present analysis. During the follow-up period, we evaluated the number of hospital admissions and inpatient days for any cause from the linked hospital care records, and the number of primary care consultations, primary care diagnostic and monitoring tests, and medication prescription items by primary care clinicians from the linked primary care records. We focused on the impact on healthcare use of the first occurrence of four cardiovascular events post-enrolment: myocardial infarction (MI), stroke, coronary revascularization (CRV), and vascular death (VD). The identification of baseline characteristics, disease events and healthcare use and the approach to missing data imputation were previously reported^11,12^.

### Statistical Models of healthcare use

The healthcare use outcomes were established over annual periods from recruitment in UKB by summing up the number of respective resource uses incurred by each participant during each year of follow-up in the study. Annual numbers of each type of healthcare use were modelled using two-part models with the first part modelling the probability of any healthcare use using logistic regression, and the second part modelling the number of non-zero healthcare use, using generalised linear model with Gamma distribution and identity link^13^. This model was chosen based on the comparison of model performance across different models (see **Supplementary section 1**).

Each model included the following pre-specified participant characteristics at entry: sex, ethnicity, quintile of Townsend deprivation index, smoking status, physical activity, diet quality, body mass index, low- and high-density lipoprotein cholesterol, serum creatinine, systolic and diastolic blood pressure, use of antihypertensive treatment and histories of diabetes mellitus, severe mental illness or cardiovascular disease. Each model included annually updated temporal histories of the first occurrences of MI, stroke, and CRV during follow-up, each of which had the following 4 categories: no event; same year (event in the annual period); 1 year ago (event in the previous annual period); and ≥2 years ago (event in an annual period two or more years ago). The annually updated participant characteristics also included current age, temporal histories of incident diabetes or cancer, and vascular and non-vascular death. We included the interactions between same year vascular death and each of the other vascular events (MI, stroke, CRV), and between the temporal history of MI and the temporal history of CRV. Cluster robust standard errors were estimated acknowledging the lack of independence between annual periods for the same participant.

Model performance was checked in deciles of predicted annual use overall and by age group and prior CVD history (**Figure S1**). The mean absolute and relative excess annual healthcare use associated with CVD events in the UKB was estimated using the estimated models and recycled prediction. This was calculated by subtracting (for absolute excess) or dividing (for relative excess) the predicted annual healthcare use in years with a CVD event by the predicted use in years without a CVD event.

### CVD microsimulation model integrated with healthcare use models

Statin effects on healthcare use were assessed by propagating the statin effects on CVD adverse events to associated healthcare use. The previously reported UK CVD microsimulation policy model^11^ was used to perform this assessment. Briefly, this decision-analytic model projects annually the first occurrence of four CVD events: MI, stroke, CRV and VD; and three key non-vascular events: incident diabetes, incident cancer and non-vascular death with the occurrence of any of these non-fatal events impacting the risks of subsequent events. The model was developed using the individual participant data of 16 large statin versus control randomised clinical trials^14^, calibrated using the UKB data^10^, and validated across categories of UKB^10^ and Whitehall II cohort^15^ participants, and against national mortality and cancer incidence rates and other published data. The model was previously used to project the event risks and survival over individuals’ remaining lifetimes (i.e. death or reaching 110 years of age) without and with statin therapy^11^, and to assess the cost-effectiveness of different statin therapies in categories of individuals^5,6^. This microsimulation model was adapted by integrating the healthcare use models (**Figure S2**), enabling the projection of healthcare use without and with statin therapy over time.

### Effects of statin therapy on healthcare use of UK Biobank participants

We assessed the lifetime effects of atorvastatin 40 mg/day on healthcare use using UKB participants’ data. We randomly sampled 10,000 UKB participants from each of the UKB subpopulations by age (40-60 and 60-70 years) and prior CVD history (without and with) at recruitment (**Table S2**) to use in the simulation of statin’s effects in the adapted CVD policy model. As in previous work^5^, effects of statin treatment on CVD events^4,16^ and adverse effects of statin^17-19^ were informed from meta-analyses of trials and cohort studies (**Table S3)**. We summarised the effects of statin therapy on cumulative healthcare use per person-year and per person over time for each of the four subpopulations by age and prior CVD history. The mean estimate was derived from base-case analysis running 500 microsimulations per individual. Uncertainty around the estimates were derived from 500 Monte-Carlo simulations capturing the uncertainty in effects of statin therapy, event risk equations and healthcare use equations related to participant characteristics and events^5,11^. (**Figure S3-S4**)

## Results

### Study population

A total of 501,807 UKB participants were included in the study, with 501,807 and 192,983 participants, contributing to hospital and primary care use analyses, respectively (**Figure S5**). Compared to participants aged 40-60 years, participants aged 60-70 years were more likely to have prior CVD history. Participants aged 60-70 and those with prior CVD history were more likely to be male, former or current smokers, with higher BMI, higher creatinine levels, with history of hypertension, diabetes, cancer, or severe mental illness (**Table 1**).

**Table 1.** Baseline characteristics of UKB study participants by age and CVD history.

|  | Age ≥40, <60 years |  | Age ≥60, ≤70 years |  |
| --- | --- | --- | --- | --- |
|  | Without CVD<br>history<br>(n = 264 237) | With CVD<br>history<br>(n = 20 390) | Without CVD<br>history<br>(n = 180 299) | With CVD<br>history<br>(n = 36 881) |
| Age (years) | 50.6 (5.6) | 52.6 (5.2) | 64.0 (2.8) | 64.8 (2.9) |
| Male | 115 144 (43.6) | 10 956 (53.7) | 79 835 (44.3) | 22 773 (61.7) |
| Ethnicity |  |  |  |  |
| White | 243 738 (92.2) | 18 726 (91.8) | 174 226 (96.6) | 35 395 (96.0) |
| Black | 5 896 (2.2) | 472 (2.3) | 1 370 (0.8) | 298 (0.8) |
| South Asian | 5 301 (2.0) | 525 (2.6) | 1 682 (0.9) | 533 (1.4) |
| Other* | 7 852 (3.0) | 527 (2.6) | 2 060 (1.1) | 435 (1.2) |
| Missing | 1 450 (0.5) | 140 (0.7) | 961 (0.5) | 220 (0.6) |
| Townsend socioeconomic deprivation (Quintile 1 = least deprived) |  |  |  |  |
| Quintile 1 | 92 873 (35.1) | 6 006 (29.5) | 73 166 (40.6) | 12 945 (35.1) |
| Quintile 2 | 51 119 (19.3) | 3 543 (17.4) | 38 092 (21.1) | 7 396 (20.1) |
| Quintile 3 | 44 130 (16.7) | 3 280 (16.1) | 28 362 (15.7) | 5 739 (15.6) |
| Quintile 4 | 40 935 (15.5) | 3 574 (17.5) | 23 423 (13.0) | 5 581 (15.1) |
| Quintile 5 | 34 783 (13.2) | 3 946 (19.4) | 17 100 (9.5) | 5 192 (14.1) |
| Missing | 397 (0.2) | 41 (0.2) | 156 (0.1) | 28 (0.1) |
| Smoking |  |  |  |  |
| Never | 155 384 (58.8) | 10 006 (49.1) | 92 912 (51.5) | 14 874 (40.3) |
| Former smoker | 75 839 (28.7) | 6 909 (33.9) | 71 942 (39.9) | 18 089 (49.0) |
| Current smoker | 31 685 (12.0) | 3 341 (16.4) | 14 294 (7.9) | 3 586 (9.7) |
| Missing | 1 329 (0.5) | 134 (0.7) | 1 151 (0.6) | 332 (0.9) |
| Physical activity |  |  |  |  |
| Low | 42 715 (16.2) | 4 031 (19.8) | 23 206 (12.9) | 6 073 (16.5) |
| Moderate | 86 706 (32.8) | 6 021 (29.5) | 59 440 (33.0) | 11 656 (31.6) |
| High | 86 555 (32.8) | 5 962 (29.2) | 58 637 (32.5) | 10 814 (29.3) |
| Missing | 48 261 (18.3) | 4 376 (21.5) | 39 016 (21.6) | 8 338 (22.6) |
| Diet quality |  |  |  |  |
| Healthy | 162 564 (61.5) | 12 015 (58.9) | 123 425 (68.5) | 23 555 (63.9) |
| Unhealthy | 95 686 (36.2) | 7 752 (38) | 53 391 (29.6) | 12 414 (33.7) |
| Missing | 5 987 (2.3) | 623 (3.1) | 3 483 (1.9) | 912 (2.5) |
| Body mass index (kg/m<sup>2</sup>) |  |  |  |  |
| <18.5 | 1 539 (0.6) | 132 (0.6) | 825 (0.5) | 121 (0.3) |
| ≥18.5, <25 | 93 352 (35.3) | 5 432 (26.6) | 55 494 (30.8) | 7 920 (21.5) |
| ≥25, <30 | 106 638 (40.4) | 7 736 (37.9) | 81 319 (45.1) | 16 138 (43.8) |
| ≥30, <35 | 43 123 (16.3) | 4 369 (21.4) | 31 273 (17.3) | 8 668 (23.5) |
| ≥35, <40 | 12 709 (4.8) | 1 613 (7.9) | 7 936 (4.4) | 2 699 (7.3) |
| ≥40 | 5 371 (2.0) | 842 (4.1) | 2 500 (1.4) | 971 (2.6) |
| Missing | 1 505 (0.6) | 266 (1.3) | 952 (0.5) | 364 (1) |
| LDL cholesterol (mmol/L) | 3.6 (0.8) | 3.3 (0.9) | 3.7 (0.9) | 3.0 (0.9) |
| HDL cholesterol (mmol/L) | 1.4 (0.4) | 1.3 (0.4) | 1.5 (0.4) | 1.3 (0.4) |
| Creatinine (umol/L) | 70.8 (14.2) | 73.6 (18.3) | 72.4 (16.2) | 78.9 (20.4) |
| Systolic BP (mmHg) | 133.4 (17.3) | 134.2 (17.8) | 144.2 (18.7) | 141.5 (19.0) |
| Diastolic BP (mmHg) | 82.2 (10.3) | 82.0 (10.5) | 82.8 (9.8) | 80.3 (10.4) |
| On antihypertensive | 26 464 (10.0) | 6 927 (34.0) | 45 461 (25.2) | 19 254 (52.2) |
|  | Without CVD<br>history<br>(n = 264 237) | With CVD<br>history<br>(n = 20 390) | Without CVD<br>history<br>(n = 180 299) | With CVD<br>history<br>(n = 36 881) |
| treatment |  |  |  |  |
| History of diabetes |  |  |  |  |
| No | 254 398 (96.3) | 18 035 (88.5) | 168 576 (93.5) | 31 066 (84.2) |
| Type 1 | 1 390 (0.5) | 470 (2.3) | 1 097 (0.6) | 908 (2.5) |
| Type 2 | 8 449 (3.2) | 1 885 (9.2) | 10 626 (5.9) | 4 907 (13.3) |
| History of cancer | 13 880 (5.3) | 1 557 (7.6) | 18 832 (10.4) | 4 302 (11.7) |
| Severe mental illness<br>history | 23 749 (9.0) | 2 920 (14.3) | 12 333 (6.8) | 3 403 (9.2) |
| History of CVD |  |  |  |  |
| No | 264 237 (100) | 0 (0) | 180 299 (100) | 0 (0) |
| MI only | 0 (0) | 623 (3.1) | 0 (0) | 1 447 (3.9) |
| Stroke only | 0 (0) | 2 030 (10.0) | 0 (0) | 3 107 (8.4) |
| PAD only | 0 (0) | 3 456 (16.9) | 0 (0) | 3 349 (9.1) |
| Other CHD only^ | 0 (0) | 10 232 (50.2) | 0 (0) | 18 737 (50.8) |
| Two or more | 0 (0) | 4 049 (19.9) | 0 (0) | 10 241 (27.8) |

BP, blood pressure; CHD, coronary heart disease; CVD, cardiovascular disease; HDL, high-density lipoprotein; LDL, low-density lipoprotein; PAD, peripheral arterial disease; SD, standard deviation. Values are mean (SD) or number (%).
\*Other ethnicity includes Chinese, Mixed, White and Black Caribbean, White and Black African, White and Asian, Any other mixed background and other ethnic group.
^Other CHD includes acute rheumatic fever, chronic rheumatic heart diseases, hypertensive heart disease, angina pectoris, other acute ischaemic heart disease, chronic ischaemic heart disease, pulmonary heart disease and other form of heart disease.

### Impact of CVD events on healthcare use

Study participants were followed for a mean duration of 7 years from recruitment. Compared to those without prior CVD, participants with CVD history were more likely to experience cardiovascular events during follow-up: MI (3.9% vs. 1.0%), stroke (3.3% vs. 0.9%), CRV (5.4% vs. 1.3%) and VD (2.5% vs. 0.4%). They also had larger annual healthcare use per person: hospital admissions (0.7 vs. 0.3), inpatient days (1.9 vs. 0.7), primary care consultations (8.3 vs. 5.1), primary care diagnostic and monitoring tests (5.8 vs. 3.0), and primary care medication prescription items (48.3 vs. 17.9) (**Table S4**). In the two-part regression models (**Table S5-S6**), MI, stroke and CRV events were associated with higher healthcare use. These excesses decreased over the two years following the events (except for medication prescription items, where the effect remained high), but longer-term higher use remained. Compared to years without an event, the relative ratio (95% CI) of hospital admissions more than two years following a CVD event ranged from 1.29 (1.22-1.36) to 1.56 (1.43-1.70), inpatient days 1.18 (1.10-1.27) to 2.95 (2.46-3.43), primary care consultations from 1.18 (1.13-1.22) to 1.31 (1.25-1.36), primary care diagnostic and monitoring tests from 1.36 (1.29-1.42) to 1.61 (1.51-1.71), and primary care prescription items from 2.10 (1.99-2.20) to 2.42 (2.18-2.66). VD was associated with higher use of hospital services but lower use of primary care services in the year of death. (**Figure 1, Table S7**)

**Figure 1.**
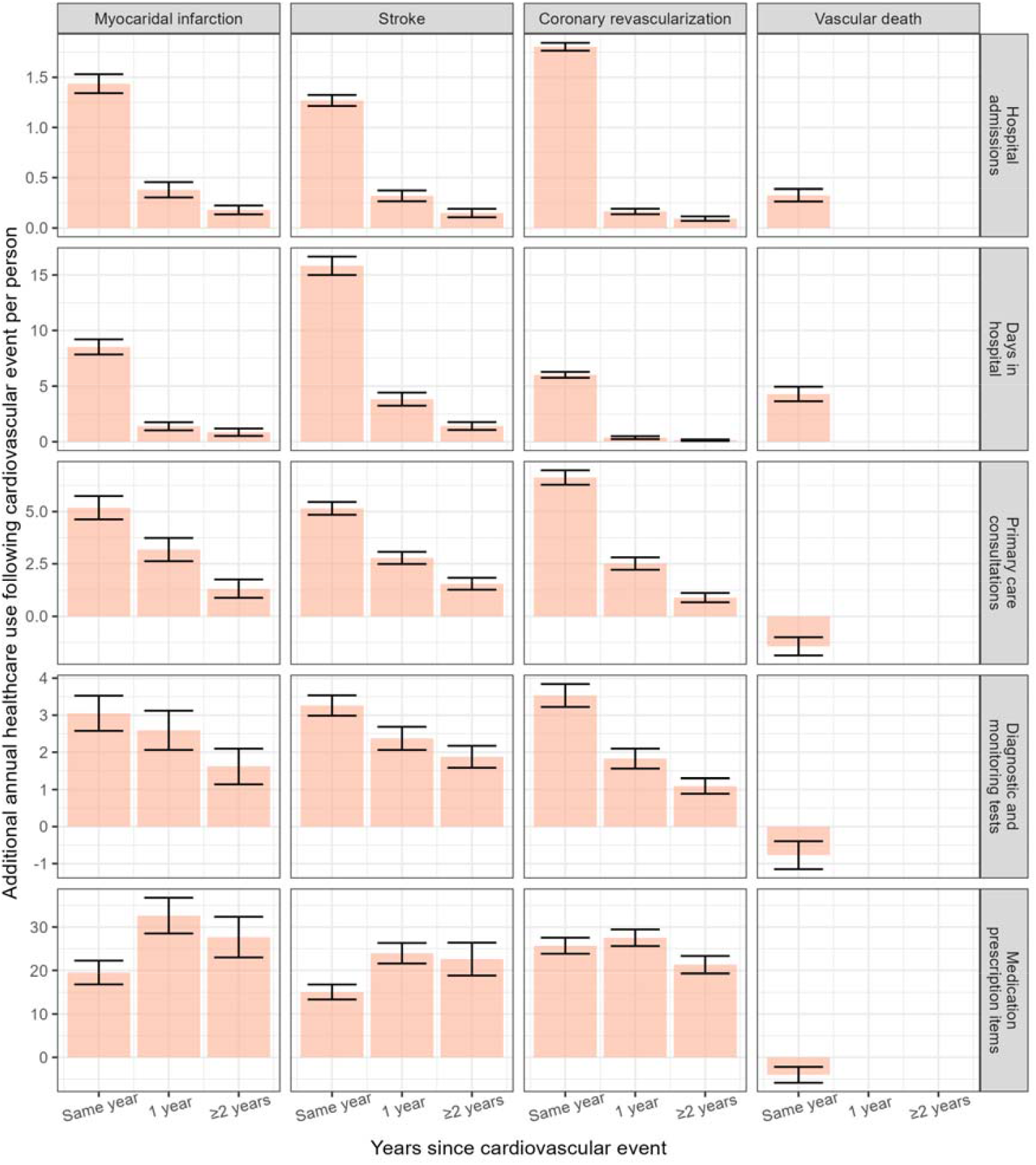
Estimated temporal impacts of cardiovascular events on healthcare services. Bars represent estimated means and error bars represent the 95% confidence intervals of the means.

### Burden of illness in the absence of statin treatment

In the absence of statin treatment, the predicted 10-year risk of new major vascular event (MVE, defined as MI, stroke, CRV, or VD) ranged from 3.3% to 6.9% (for individuals aged 40-60 to 60-70 respectively) in those without CVD history, and from 13.8% to 21.4% in those with prior CVD history. Over a lifetime, these risks increased to 35.6%-38.2% and 52.7%-53.5%, respectively. The predicted remaining life expectancy was 35.2 and 26.3 years for participants age 40-60 years and 60-70 years without prior CVD history, decreasing to 27.4 and 20.8 years respectively in those with prior CVD history (**Table S8**). Estimated rate of healthcare use increased over time and was consistently higher among older individuals and those with CVD history. Per person-year, over the first 10 years, mean number of hospital admissions ranged from 0.29 (aged 40-60 without CVD history) to 0.85 (aged 60-70 with CVD history), rising to 0.52-1.06 over lifetime. Hospital inpatient days increased from 0.65-2.49 to 1.59-3.57, primary care consultations from 4.66-8.90 to 6.04-9.75, primary care diagnostic and monitoring tests from 2.76-6.51 to 3.94-7.32, and primary care medication prescription items from 14.7-52.1 to 24.2-59.7. (**Figure 2, Table S9**)

**Figure 2.**
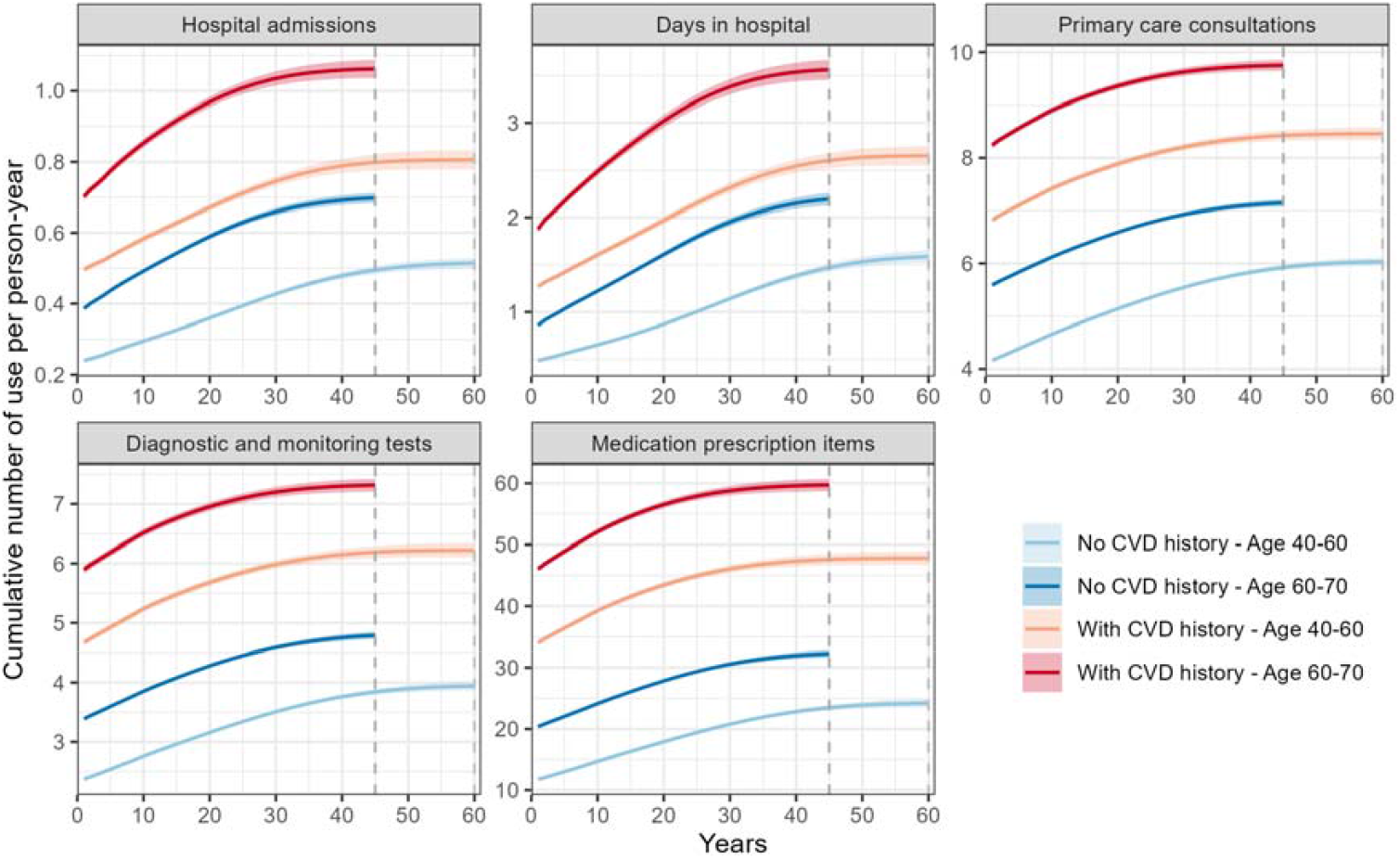
Predicted cumulative healthcare use without statin treatment overtime. Lines represent the mean estimates and shaded areas represent the 95% confidence intervals of the means. Assuming all participants died at age 110 years, the summary ends in year 60 (light grey dashed line) for people aged 40-60 and in year 45 for people aged 60-70 (grey dashed line).

### Impact of statin treatment on hospital admissions and primary care use

Over longer time horizons, statin treatment was associated with larger reductions in the proportion of people with new MVE, ranging from 1.2%-2.5% and 4.8%-7.1% in people without and with CVD history respectively over 10 years to 7.5%-8.9% and 10.2%-11.5%, respectively, over lifetime. These effects were paralleled by increasing gains in life expectancy ranging from 0.005-0.015 and 0.039-0.062 years per person over 10 years among people without and with prior CVD history, respectively, to 0.64-0.71 and 0.80-0.94 over lifetime. (**Figure 3, Table S9**)

**Figure 3.**
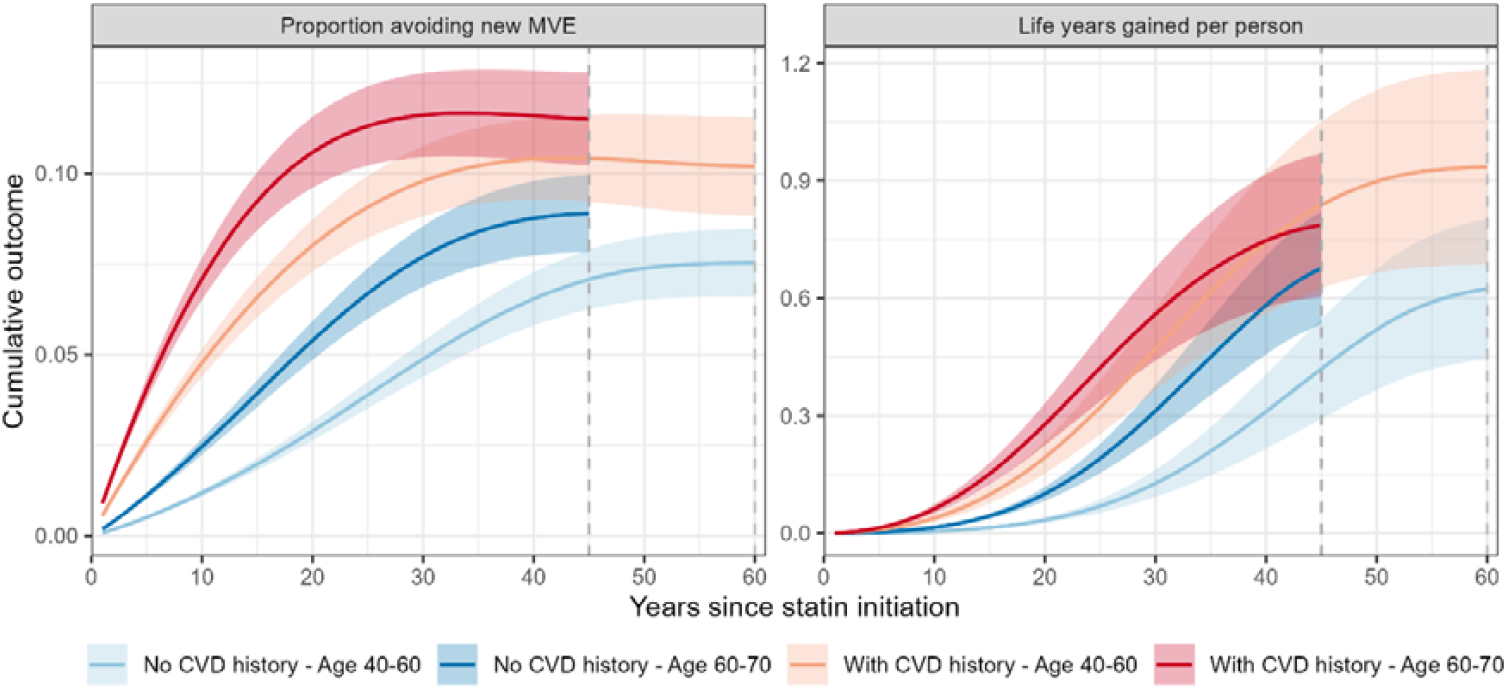
Effects of statin treatment on major vascular events and survival over time. Lines represents the mean estimates and shaded areas represent the 95% confidence intervals of the means. MVE: major vascular event, including myocardial infarction, stroke, coronary revascularization and vascular death. Assuming all participants died at age 110 years, the summary ends in year 60 for people aged 40-60 and in year 45 for people aged 60-70.

The joint effects of statin treatment in reducing the risk of CVD events and increasing life expectancy led to large reductions in healthcare use in earlier years after statin initiation followed by an increased longer-term use. The estimated time from statin initiation to neutral net effect on cumulative healthcare use occurred earliest for primary care tests and consultations [ranging from 9 (7-12) to 17 (9-25) years] respectively, followed by primary care medication prescription items and hospital admissions [22 (17-28) to 38 (28-48) years] respectively, and hospital inpatient days [40 (30-47) to 51 (43-62) years]. These transitions occurred earlier in older individuals and those with prior CVD history. Over lifetime, statin treatment led to an estimated per-person increase of 0.33 (0.17, 0.48) to 0.60 (0.33, 0.88) hospital admissions, 0.29 (-0.35, 0.93) to 0.89 (-0.14, 1.92) hospital admission days, 4.93 (3.48, 6.38) to 8.35 (5.94, 10.76) consultations, 3.70 (2.57, 4.82) to 6.39 (4.51, 8.27) tests, and 15.4 (6.2. 24.5) to 33.6 (17.8, 49.3) prescription items. (**Figure 4, Table S10**) Unlike healthcare use per person, healthcare use per person-year remained lower over lifetime for hospital admissions [0.0000002 (-0.002, 0.002) to -0.014 (-0.019, -0.009)] and hospital inpatient days [-0.016 (-0.030, - 0.002) to -0.115 (-0.150, -0.081)], and primary care prescription items [-0.002 (-0.196, 0.191) to – 0.927 (-1.252, -0.602)]. (**Figure 4, Table S9**)

**Figure 4.**
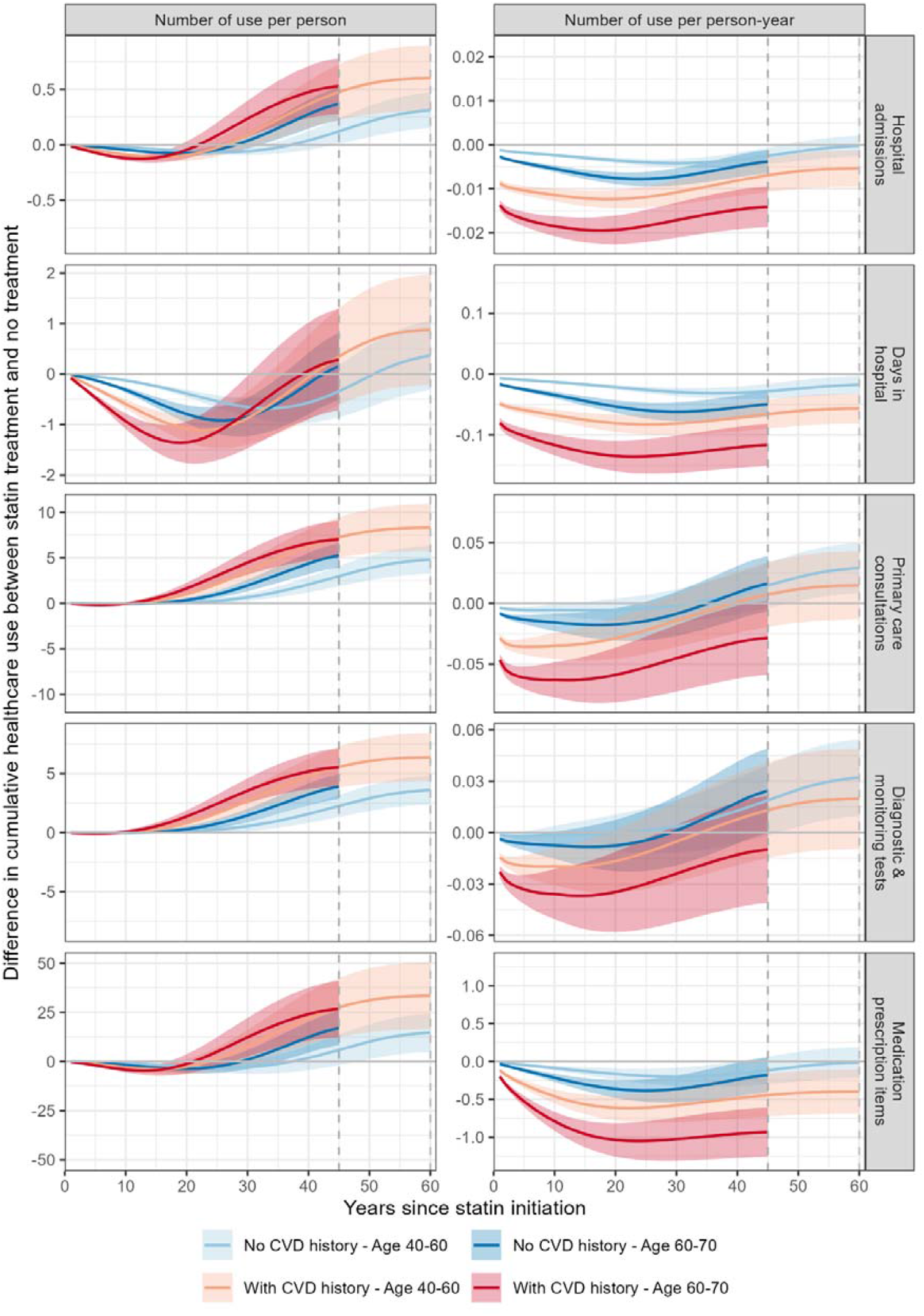
Effects of statin treatment on cumulative healthcare use. Lines represent the mean estimates and shaded areas represent the 95% confidence intervals of the means. Assuming all participants died at age 110 years, the summary ends in year 60 for people aged 40-60 and in year 45 for people aged 60-70.

## Discussion

This study provides novel insights into the impact of statin therapy on healthcare use over time. We report that MI, stroke, and CRV events were associated with marked increases in healthcare use in the first two years post events which declined but remained elevated thereafter, with persistently high demand for medications. Statin treatment resulted in increased survival and lower rates of hospital admissions, inpatient days and primary care prescriptions per person-year over lifetime, with larger reductions for older adults and those with a prior CVD history. Over lifetime, however, per person net healthcare use was greater among statin users. Transitions to greater net healthcare use occurred sooner for primary care consultations and tests (10–20 years after statin initiation) and later for hospital inpatient days (40–50 years after statin initiation), with earlier transitions observed in older individuals and those with a prior history of CVD.

Our findings align with McConnachie *et al*.^20^, which showed that allocation to statin over five years in the WOSCOPS study significantly reduced cardiovascular-related hospital admissions and inpatient days, with a modest increase in admissions for other causes over 15 years. We extend this evidence by assessing estimated statin effects with longer duration of statin treatment, over a longer timeframe, across a broader population (including women and individuals with prior CVD), and on additional healthcare resources such as primary care services. In contrast, Cooke *et al*. found no association between statin use and reductions in re-hospitalizations or physician visits.^21^ However, their analysis spanned only about two years — likely too short to detect differences. In addition, they focused on statin use in 1997-2001, a period dominated by less potent statin formulations.

Despite robust evidence for statins’ efficacy and safety, millions of statin eligible people in the UK remain untreated,^22^ and adherence is often poor even among individuals with prior CVD^23^. Programs aimed at improving adherence to statin therapy, including pharmacist interventions^24^, mobile phone text messaging^25^, and financial incentives^26^, have limited evidence for effectiveness and cost-effectiveness. Our study provides new evidence that statin treatment reduces average rate of healthcare use, particularly hospital admissions, broadening their value beyond clinical benefits. Communicating the benefits of preventive interventions such as statin therapy on reducing hospital admission in addition to reducing risk of CVD events — could improve public awareness of the broader consequences of CVD and benefits of prevention and further incentivise heathier lifestyles and adherence to preventive measures^3,27^.

We also show how statins impact long term healthcare use. Among statin treated individuals, healthcare use is progressively lower after statin initiation, levelling off after the first decade and transitioning into increased use after the second decade — earlier for primary care services and in older individuals or those with prior CVD history. The increased survival with statin treatment contributed to increased healthcare use in later years as a result of ageing and morbidity such as cancer and diabetes (**Figure S6**) among survivors. These insights can inform healthcare resource planning. While our analysis is UK-based, the findings are likely relevant to other settings, including resource-limited setting.^8^

The key strengths of the study include the use of the large-scale, population-based UKB cohort, which enabled a comprehensive assessment of healthcare use linked to CVD events across a diverse, real-world population. The integration of the detailed healthcare use data into a validated CVD microsimulation model allowed for a robust estimation of statin impact on healthcare use over time— addressing a gap in the literature on healthcare use implications of statin treatment and illustrating broader impact of preventive therapies. A study limitation is the lack of data availability for hospital outpatient, laboratory services, emergency department care, and social care use; further research to study statin effects on the broader range of healthcare and social care services will be helpful. Finally, the UK Biobank cohort is not representative of the general UK population, with reduced social diversity and a healthy volunteer effect. However, we present findings by two key factors, age and prior CVD history, which we hope improves generalizability.

In summary, our study quantifies the increased healthcare use following CVD events, and statins’ longer term effects on healthcare use. While statins reduce healthcare use in earlier years, extended life expectancy and ageing lead to increased healthcare use in survivors in later years, especially for primary care services and among older people or people with a prior CVD history at statin initiation. These dynamics underscore the need for long-term planning in healthcare resource allocation, with a shift from acute CVD care toward sustained preventive and chronic care services.

## Declaration statement

### Data availability statement

Data may be obtained from a third party and are not publicly available. The datasets used in the current study are available from UK Biobank (https://www.ukbiobank.ac.uk/). Researchers can apply to use the UK Biobank resource.

### Rights retention statement: author accepted manuscript

For the purpose of open access, the authors have applied a Creative Commons Attribution (CC BY) licence to any Author Accepted Manuscript version arising.

## Acknowledgements

This research has been conducted using data from UK Biobank, a major biomedical database www.ukbiobank.ac.uk. We thank all the participants, staff and other contributors to the resource.

## Authors’ contributions

JZ & BM developed the concept for these analyses. JZ & BM had full access to all study data and take responsibility for its integrity and analysis. JZ wrote the first draft of the manuscript with support from BM. All authors contributed to interpretation of the results and manuscript revision.

## References

1. Vaduganathan M, Mensah George A, Turco Justine V, Fuster V, Roth Gregory A. The Global Burden of Cardiovascular Diseases and Risk. JACC 2022; 80(25): 2361–71.

2. British Heart Foundation. BHF UK CVD Factsheet 2025. https://wwwbhforguk/-/media/files/for-professionals/research/heart-statistics/bhf-cvd-statistics-uk-factsheetpdf (Accessed on 2025/06/15) 2025.

3. National Institute for Health and Care Excellence. Cardiovascular disease: risk assessment and reduction, including lipid modification. Clinical Guideline; 2023 [cited 12/11/2024] Available from: https://wwwniceorguk/guidance/ng238.

4. Cholesterol Treatment Trialists C, Mihaylova B, Emberson J, et al. The effects of lowering LDL cholesterol with statin therapy in people at low risk of vascular disease: meta-analysis of individual data from 27 randomised trials. Lancet 2012; 380(9841): 581–90.

5. Mihaylova B, Wu R, Zhou J, et al. Lifetime effects and cost-effectiveness of standard and higher-intensity statin therapy across population categories in the UK: a microsimulation modelling study. Lancet Reg Health Eur 2024; 40: 100887.

6. Mihaylova B, Wu R, Zhou J, et al. Lifetime effects and cost-effectiveness of statin therapy for older people in the United Kingdom: a modelling study. Heart 2024; 110(21): 1277–85.

7. Cholesterol Treatment Trialists C, Baigent C, Blackwell L, et al. Efficacy and safety of more intensive lowering of LDL cholesterol: a meta-analysis of data from 170,000 participants in 26 randomised trials. Lancet 2010; 376(9753): 1670–81.

8. National Academies of Sciences, Engineering, and Medicine; Health and Medicine Division; Board on Health Care Services; Committee on Health Care Utilization and Adults with Disabilities. Health-Care Utilization as a Proxy in Disability Determination. Washington (DC): National Academies Press (US); 2018 Mar 1. 2, Factors That Affect Health-Care Utilization. Available from: https://www.ncbi.nlm.nih.gov/books/NBK500097/.

9. Taber JM, Leyva B, Persoskie A. Why do people avoid medical care? A qualitative study using national data. J Gen Intern Med 2015; 30(3): 290–7.

10. Sudlow C, Gallacher J, Allen N, et al. UK biobank: an open access resource for identifying the causes of a wide range of complex diseases of middle and old age. PLoS Med 2015; 12(3): e1001779.

11. Wu R, Williams C, Zhou J, et al. Long-term cardiovascular risks and the impact of statin treatment on socioeconomic inequalities: a microsimulation model. Br J Gen Pract 2024; 74(740): e189–e98.

12. Zhou J, Wu R, Williams C, et al. Prediction Models for Individual-Level Healthcare Costs Associated with Cardiovascular Events in the UK. Pharmacoeconomics 2023.

13. Zhou J, Williams C, Keng MJ, Wu R, Mihaylova B. Estimating Costs Associated with Disease Model States Using Generalized Linear Models: A Tutorial. Pharmacoeconomics 2024; 42(3): 261-73.

14. Protocol for a prospective collaborative overview of all current and planned randomized trials of cholesterol treatment regimens. Cholesterol Treatment Trialists’ (CTT) Collaboration. Am J Cardiol 1995; 75(16): 1130–4.

15. Marmot M, Brunner E. Cohort Profile: the Whitehall II study. Int J Epidemiol 2005; 34(2): 251–6.

16. Law MR, Wald NJ, Rudnicka AR. Quantifying effect of statins on low density lipoprotein cholesterol, ischaemic heart disease, and stroke: systematic review and meta-analysis. BMJ 2003; 326(7404): 1423.

17. Law M, Rudnicka AR. Statin safety: a systematic review. Am J Cardiol 2006; 97(8A): 52C–60C.

18. Preiss D, Seshasai SR, Welsh P, et al. Risk of incident diabetes with intensive-dose compared with moderate-dose statin therapy: a meta-analysis. JAMA 2011; 305(24): 2556–64.

19. Sattar N, Preiss D, Murray HM, et al. Statins and risk of incident diabetes: a collaborative meta-analysis of randomised statin trials. Lancet 2010; 375(9716): 735–42.

20. McConnachie A, Walker A, Robertson M, et al. Long-term impact on healthcare resource utilization of statin treatment, and its cost effectiveness in the primary prevention of cardiovascular disease: a record linkage study. Eur Heart J 2014; 35(5): 290–8.

21. Cooke CA, Kirkland SA, Sketris IS, Cox J. The impact of statins on health services utilization and mortality in older adults discharged from hospital with ischemic heart disease: a cohort study. BMC Health Serv Res 2009; 9: 198.

22. Ueda P, Lung TW, Clarke P, Danaei G. Application of the 2014 NICE cholesterol guidelines in the English population: a cross-sectional analysis. Br J Gen Pract 2017; 67(662): e598–e608.

23. Thalmann I, Preiss D, Schlackow I, Gray A, Mihaylova B. Population-wide cohort study of statin use for the secondary cardiovascular disease prevention in Scotland in 2009-2017. Heart 2023; 109(5): 388–95.

24. Eussen SR, van der Elst ME, Klungel OH, et al. A pharmaceutical care program to improve adherence to statin therapy: a randomized controlled trial. Ann Pharmacother 2010; 44(12): 1905–13.

25. Redfern J, Tu Q, Hyun K, et al. Mobile phone text messaging for medication adherence in secondary prevention of cardiovascular disease. Cochrane Database Syst Rev 2024; 3(3): Cd011851.

26. Barankay I, Reese PP, Putt ME, et al. Effect of Patient Financial Incentives on Statin Adherence and Lipid Control: A Randomized Clinical Trial. JAMA Netw Open 2020; 3(10): e2019429.

27. Duffy EY, Ashen D, Blumenthal RS, et al. Communication approaches to enhance patient motivation and adherence in cardiovascular disease prevention. Clin Cardiol 2021; 44(9): 1199–207.

