## Supplementary materials for "Effects of statin treatment on primary and hospital care use: a microsimulation model"

### Supplementary methods

***Supplementary section 1. Comparison of models of healthcare resource use.***

A number of models of annual use of healthcare resources (i.e. hospital admissions, days in hospital, primary care consultations, diagnostic and monitoring tests and medication prescription items) were compared, including three one-part models, five two-part models, two hurdle models and two zero-inflated models. Two-part, hurdle and zero-inflated models estimate two models, with part 1 modelling the probability of any resource use, and part 2 modelling the quantity used conditional on use. Table SS1 summarises key differences between these types of models and Table SS2 specifies the candidate models studied.

**Table SS1. Two-part, hurdle and zero-inflated models**

| **Type of model** | **Relationship between Part 1 and Part 2** | **Estimation of Part 1 and Part 2** |
|---|---|---|
| Two-part model | Independent | Separate |
| Hurdle model | Independent | Joint |
| Zero-inflated model | Dependent | Joint |

**Table SS2. Different models compared for modelling healthcare resource use**

| **Type** | **GLM in one-part model or part 2 in models with two parts** |
|---|---|
| One-part models | 1. Poisson – LOG (1P-POI-LOG)<br>2. Negative binomial – LOG (1P-NB-LOG)<br>3. Gamma – Identity (1P-GAM-ID) |
| Two-part models* | 4. Poisson-Identity (2P-POI-ID)<br>5. Poisson-LOG (2P-POI-LOG)<br>6. Negative binomial – LOG (2P-NB-LOG)<br>7. Gamma-Identity (2P-GAM-ID)<br>8. Gamma-LOG (2P-GAM-LOG) |
| Hurdle models* | 9. Poisson-LOG (HD-POI-LOG)<br>10. Negative binomial-LOG (HD-NB-LOG) |
| Zero-inflated models* | 11. Poisson-LOG (ZI-POI-LOG)<br>12. Negative binomial-LOG (ZI-NB-LOG) |

* First part models the probability of any resource use using a logistic regression.

The better performing models were those with smaller mean error (difference between predicted vs. observed annual resource use across all participant-years) in most deciles of predicted annual resource use (**Figure SS1**). For each type of model, the best performing models across all healthcare resource use outcomes were:

- One-part models: Gamma-Identity GLM;
- Two-part models: Poison-LOG and Gamma-Identity;
- Hurdle models: Poisson-LOG;
- Zero-inflated models:  Poisson-LOG.

The comparison of the mean errors by decile of predicted annual resource use in population categories by age and history of CVD across these five models indicated the following **two-part models** as the best performing across all these population categories (**Figure SS2).**

- **Part 1**: logistic regression modelling the probability of incurring any resource use;
- **Part 2**: Gamma-identity GLM modelling the quantity of resource use conditional on any use.

These models were used further in the study.

**Figure SS1. Model performance by deciles of predicted annual healthcare resource use in the study population**

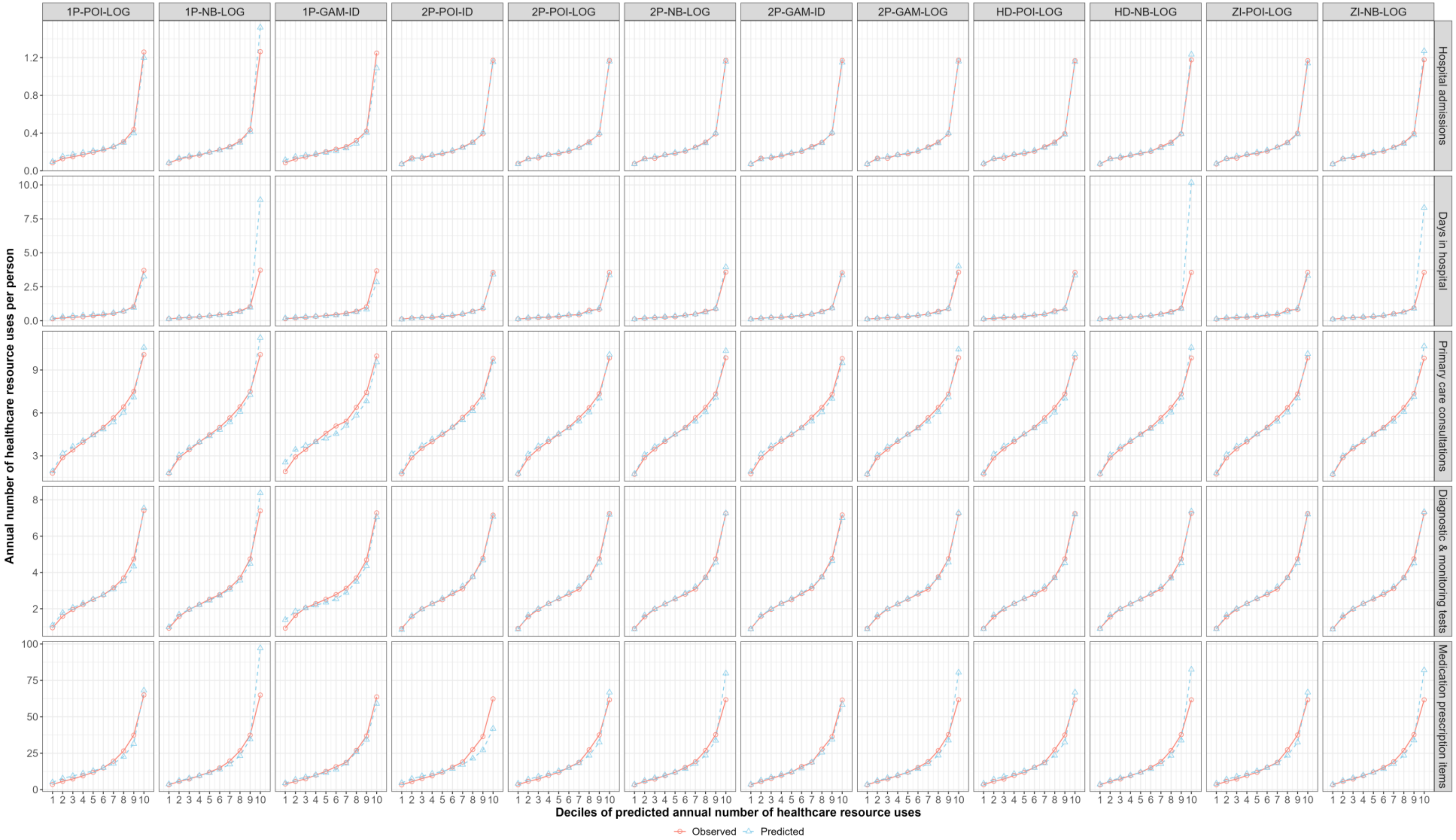

We only described the second part of the two-part, hurdle, and zero-inflated model (all part 1 is logistic regression). 1P = one-part model; 2P = Two-part model; HD = hurdle model; ZI = zero-inflated model; POI = Poisson; NB = Negative Binomial; Gam = Gamma; ID = Identity; LOG = natural logarithm.

**Figure SS2. Model performance by deciles of predicted outcomes in categories of study population**

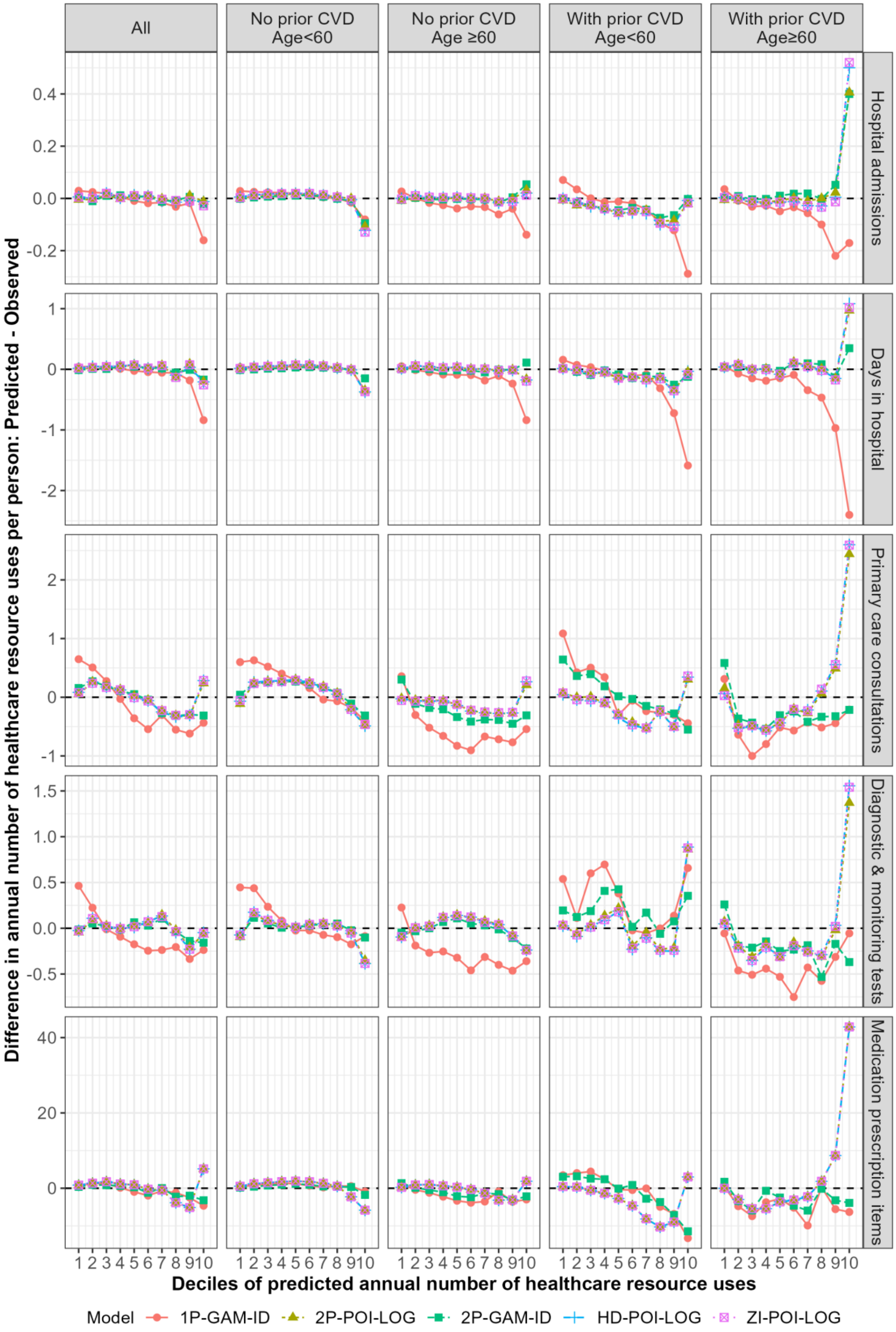

We only described the second part of the two-part, hurdle, and zero-inflated model (all part 1 is logistic regression).1P = one-part; 2P=Two-part; HD=hurdle; ZI = zero-inflated; POI = Poisson; Gam = Gamma; ID = Identity; LOG = natural logarithm;

### Supplementary tables and figures

***Supplementary table 1. Baseline characteristics of participants included in hospital care use analysis contributing and not contributing to primary care use analysis.***

| | **Included in primary care use analysis** | | **Excluded from primary care use analysis** | |
|---|---|---|---|---|
| | **Without CVD history (n = 168 205)** | **With CVD history (n = 24 778)** | **Without CVD history (n = 276 331)** | **With CVD history (n = 32 493)** |
| Age (years) | 56.1 (8.0) | 60.3 (7.1) | 56.0 (8.1) | 60.5 (7.0) |
| Male | 73 573 (43.7) | 14 084 (56.8) | 121 406 (43.9) | 19 645 (60.5) |
| Ethnicity | | | | |
| White | 159 517 (94.8) | 23 570 (95.1) | 258 447 (93.5) | 30 551 (94) |
| Black | 1 851 (1.1) | 222 (0.9) | 5 415 (2) | 548 (1.7) |
| South Asian | 2 905 (1.7) | 495 (2) | 4 078 (1.5) | 563 (1.7) |
| Other* | 3 170 (1.9) | 355 (1.4) | 6 742 (2.4) | 607 (1.9) |
| Missing | 762 (0.5) | 136 (0.5) | 1 649 (0.6) | 224 (0.7) |
| Townsend socioeconomic deprivation | | | | |
| Quintile 1 (least deprived) | 52 210 (31) | 6 571 (26.5) | 86 579 (31.3) | 8 438 (26) |
| Quintile 2 | 37 736 (22.4) | 5 223 (21.1) | 60 414 (21.9) | 6 499 (20) |
| Quintile 3 | 29 506 (17.5) | 4 397 (17.7) | 46 164 (16.7) | 5 476 (16.9) |
| Quintile 4 | 25 853 (15.4) | 4 132 (16.7) | 42 519 (15.4) | 5 377 (16.5) |
| Quintile 5 | 18 554 (11) | 3 809 (15.4) | 33 752 (12.2) | 5 882 (18.1) |
| Missing | 4 346 (2.6) | 646 (2.6) | 6 903 (2.5) | 821 (2.5) |
| Smoking | | | | |
| Never | 94 763 (56.3) | 11 069 (44.7) | 153 533 (55.6) | 13 811 (42.5) |
| Former smoker | 55 607 (33.1) | 10 654 (43) | 92 174 (33.4) | 14 344 (44.1) |
| Current smoker | 17 016 (10.1) | 2 874 (11.6) | 28 963 (10.5) | 4 053 (12.5) |
| Missing | 819 (0.5) | 181 (0.7) | 1 661 (0.6) | 285 (0.9) |
| Physical activity | | | | |
| Low | 24 777 (14.7) | 4 265 (17.2) | 41 144 (14.9) | 5 839 (18) |
| Moderate | 55 199 (32.8) | 7 676 (31) | 90 947 (32.9) | 10 001 (30.8) |
| High | 55 680 (33.1) | 7 468 (30.1) | 89 512 (32.4) | 9 308 (28.6) |
| Missing | 32 549 (19.4) | 5 369 (21.7) | 54 728 (19.8) | 7 345 (22.6) |
| Diet quality | | | | |
| Healthy | 108 313 (64.4) | 15 553 (62.8) | 177 676 (64.3) | 20 017 (61.6) |
| Unhealthy | 56 945 (33.9) | 8 643 (34.9) | 92 132 (33.3) | 11 523 (35.5) |
| Missing | 2 947 (1.8) | 582 (2.3) | 6 523 (2.4) | 953 (2.9) |
| Body mass index (kg/m$^2$) | | | | |
| <18.5 | 840 (0.5) | 128 (0.5) | 1 524 (0.6) | 125 (0.4) |
| ≥18.5, <25 | 55 226 (32.8) | 6 035 (24.4) | 93 620 (33.9) | 7 317 (22.5) |
| ≥25, <30 | 71 515 (42.5) | 10 344 (41.7) | 116 442 (42.1) | 13 530 (41.6) |
| ≥30, <35 | 28 779 (17.1) | 5 485 (22.1) | 45 617 (16.5) | 7 552 (23.2) |
| ≥35, <40 | 7 949 (4.7) | 1 802 (7.3) | 12 696 (4.6) | 2 510 (7.7) |
| ≥40 | 2 990 (1.8) | 733 (3) | 4 881 (1.8) | 1 080 (3.3) |
| Missing | 906 (0.5) | 251 (1) | 1 551 (0.6) | 379 (1.2) |
| LDL cholesterol (mmol/L) | 3.6 (0.9) | 3.1 (0.9) | 3.6 (0.8) | 3.1 (0.9) |
| HDL cholesterol (mmol/L) | 1.5 (0.4) | 1.3 (0.4) | 1.5 (0.4) | 1.3 (0.4) |
| Creatinine (umol/L) | 71.5 (15.1) | 76.5 (19.5) | 71.5 (15.0) | 77.4 (20.1) |

| | Included in primary care use analysis | | Excluded from primary care use analysis | |
|---|---|---|---|---|
| | **Without CVD history (n = 168 205)** | **With CVD history (n = 24 778)** | **Without CVD history (n = 276 331)** | **With CVD history (n = 32 493)** |
| Systolic blood pressure (mmHg) | 138.2 (18.7) | 139.1 (19.0) | 137.5 (18.6) | 138.7 (18.9) |
| Diastolic blood pressure (mmHg) | 82.6 (10.1) | 81.0 (10.4) | 82.3 (10.1) | 80.8 (10.5) |
| On antihypertensive treatment | 27 240 (16.2) | 10 900 (44) | 44 685 (16.2) | 15 281 (47) |
| History of diabetes | | | | |
| Type 1 | 926 (0.6) | 558 (2.3) | 1 561 (0.6) | 820 (2.5) |
| Type 2 | 7 134 (4.2) | 2 694 (10.9) | 11 941 (4.3) | 4 098 (12.6) |
| History of cancer | 12 221 (7.3) | 2 420 (9.8) | 20 491 (7.4) | 3 439 (10.6) |
| Severe mental illness history | 17 549 (10.4) | 3 374 (13.6) | 18 533 (6.7) | 2 949 (9.1) |
| History of CVD | | | | |
| No | 168 205 (100) | 0 (0) | 276 331 (100) | 0 (0) |
| MI only | 0 (0) | 776 (3.1) | 0 (0) | 1 294 (4) |
| Stroke only | 0 (0) | 1 991 (8) | 0 (0) | 3 146 (9.7) |
| PAD only | 0 (0) | 3 473 (14) | 0 (0) | 3 332 (10.3) |
| Other CHD only^ | 0 (0) | 12 642 (51) | 0 (0) | 16 327 (50.2) |
| Two or more | 0 (0) | 5 896 (23.8) | 0 (0) | 8 394 (25.8) |

CHD, coronary heart disease; CVD, cardiovascular disease; HDL, high density lipoprotein; LDL, low density lipoprotein; PAD, peripheral arterial disease; SD, standard deviation.

Values are mean (SD) or number (%).

*Other ethnicity includes Chinese, Mixed, White and Black Caribbean, White and Black African, White and Asian, Any other mixed background and other ethnic group.

^Other CHD includes acute rheumatic fever, chronic rheumatic heart diseases, hypertensive heart disease, angina pectoris, other acute ischaemic heart disease, chronic ischaemic heart disease, pulmonary heart disease and other form of heart disease.

***Supplementary figure 1. Performance of the selected two-part model (P1: logistic regression; P2: Gamma-Identity GLM) of healthcare resource use***

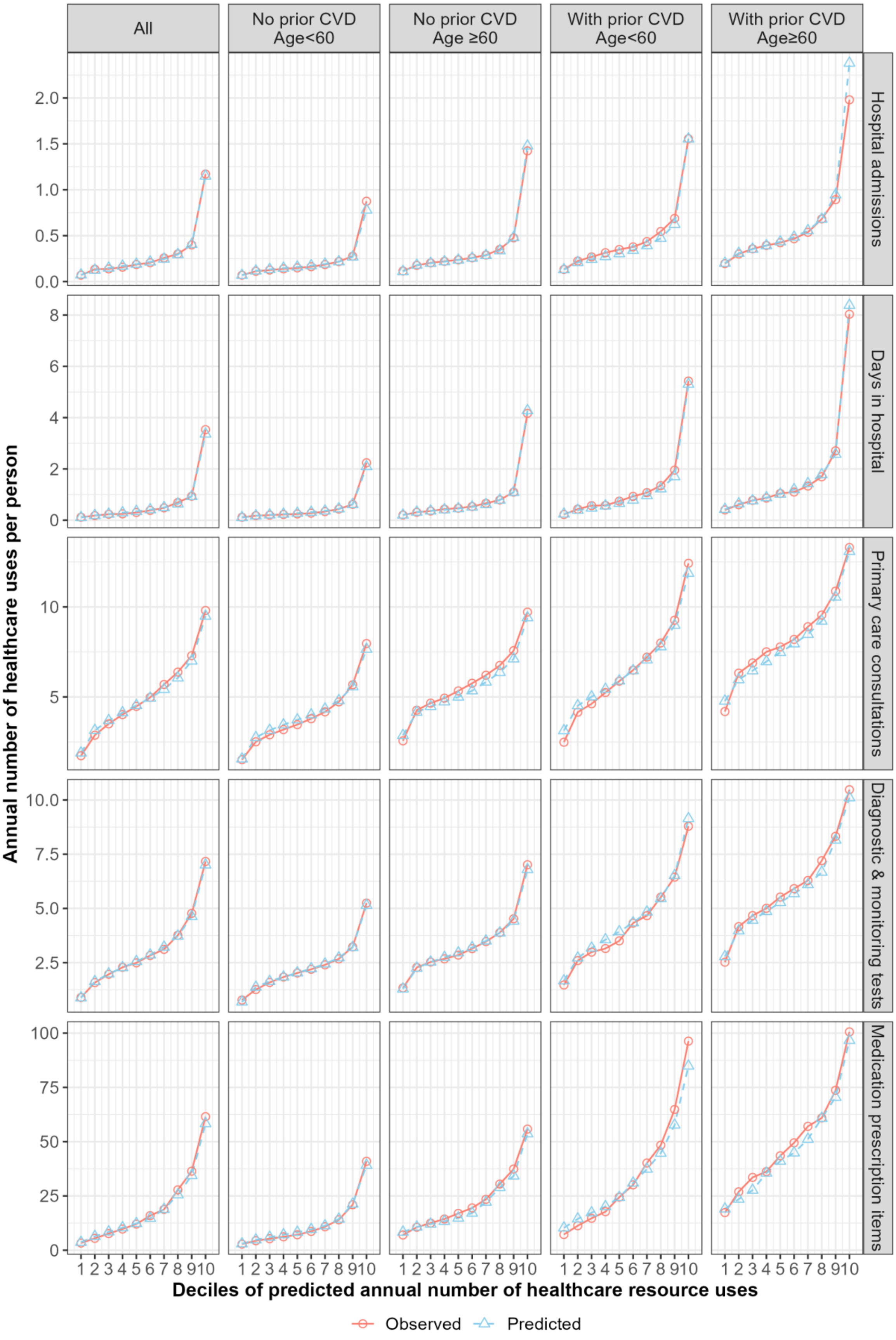

*Supplementary figure 2. Schematic of the CVD microsimulation model and integrated healthcare use models*

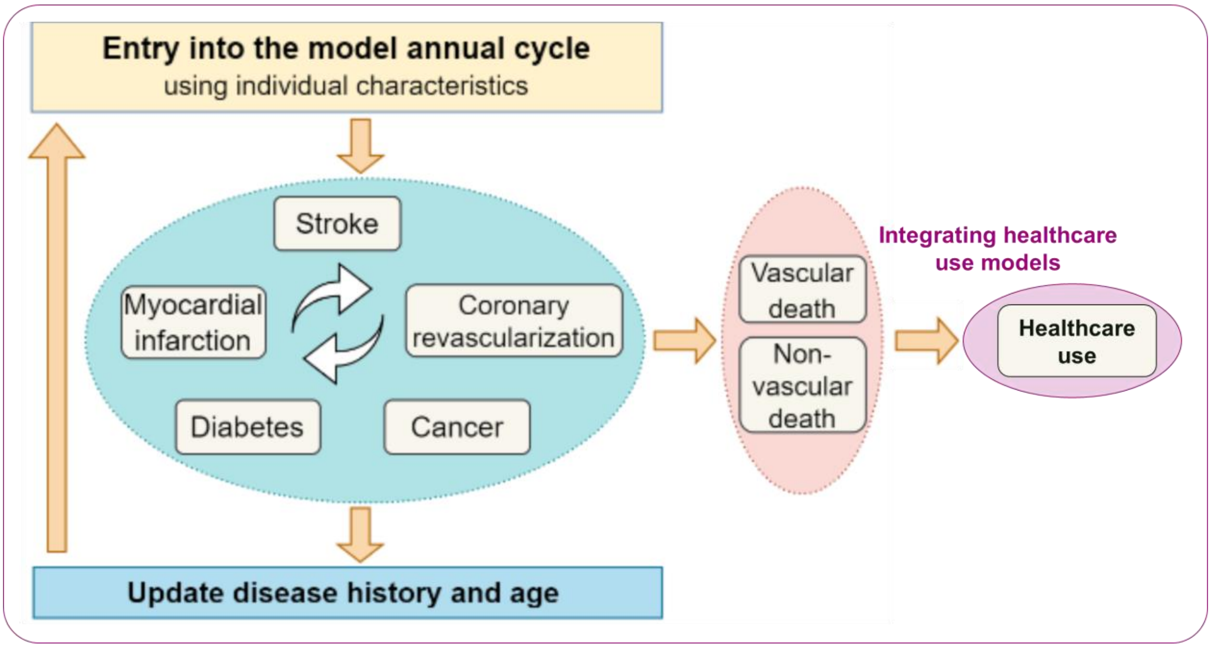

***Supplementary table 2. Baseline characteristics of the random samples of participants contributing to the assessment of statin's effects on healthcare resource use, by age and history of CVD***

| | **Age ≥40, <60 years** | | **Age ≥60, ≤70 years** | |
|---|---|---|---|---|
| | **Without CVD history (n = 10 000)** | **With CVD history (n = 10 000)** | **Without CVD history (n = 10 000)** | **With CVD history (n = 10 000)** |
| Age (years) | 50.6 (5.6) | 52.5 (5.2) | 64.0 (2.8) | 64.8 (2.9) |
| Male | 4 339 (43.4) | 5 304 (53.0) | 4 430 (44.3) | 6 100 (61.0) |
| Ethnicity | | | | |
| White | 9 279 (92.8) | 9 272 (92.7) | 9 726 (97.3) | 9 663 (96.6) |
| Black | 225 (2.2) | 243 (2.4) | 77 (0.8) | 73 (0.7) |
| South Asian | 213 (2.1) | 240 (2.4) | 86 (0.9) | 146 (1.5) |
| Other* | 283 (2.8) | 245 (2.5) | 111 (1.1) | 118 (1.2) |
| Townsend socioeconomic deprivation (Quintile 1 = least deprived) | | | | |
| Quintile 1 | 3 473 (34.7) | 2 997 (30.0) | 4 055 (40.6) | 3 551 (35.5) |
| Quintile 2 | 1 960 (19.6) | 1 755 (17.5) | 2 095 (21.0) | 1 995 (20.0) |
| Quintile 3 | 1 698 (17.0) | 1 600 (16.0) | 1 576 (15.8) | 1 482 (14.8) |
| Quintile 4 | 1 582 (15.8) | 1 717 (17.2) | 1 325 (13.2) | 1 500 (15.0) |
| Quintile 5 | 1 287 (12.9) | 1 931 (19.3) | 949 (9.5) | 1 472 (14.7) |
| Smoking | | | | |
| Never | 5 908 (59.1) | 4 922 (49.2) | 5 204 (52.0) | 4 154 (41.5) |
| Former smoker | 2 911 (29.1) | 3 443 (34.4) | 4 006 (40.1) | 4 874 (48.7) |
| Current smoker | 1 181 (11.8) | 1 635 (16.4) | 790 (7.9) | 972 (9.7) |
| Physical activity | | | | |
| Low | 1 579 (15.8) | 2 024 (20.2) | 1 313 (13.1) | 1 675 (16.8) |
| Moderate | 3 273 (32.7) | 2 958 (29.6) | 3 302 (33.0) | 3 086 (30.9) |
| High | 3 271 (32.7) | 2 868 (28.7) | 3 212 (32.1) | 2 943 (29.4) |
| Missing | 1 877 (18.8) | 2 150 (21.5) | 2 173 (21.7) | 2 296 (23.0) |
| Unhealthy diet | 3 831 (38.3) | 4 044 (40.4) | 3 176 (31.8) | 3 612 (36.1) |
| Body mass index (kg/m$^2$) | | | | |
| <18.5 | 55 (0.5) | 60 (0.6) | 48 (0.5) | 21 (0.2) |
| ≥18.5, <25 | 3 534 (35.3) | 2 722 (27.2) | 3 136 (31.4) | 2 224 (22.2) |
| ≥25, <30 | 4 092 (40.9) | 3 974 (39.7) | 4 467 (44.7) | 4 372 (43.7) |
| ≥30, <35 | 1 644 (16.4) | 2 080 (20.8) | 1 761 (17.6) | 2 396 (24.0) |
| ≥35, <40 | 464 (4.6) | 755 (7.6) | 459 (4.6) | 717 (7.2) |
| ≥40 | 211 (2.1) | 409 (4.1) | 129 (1.3) | 270 (2.7) |
| LDL cholesterol (mmol/L) | 3.6 (0.8) | 3.3 (0.9) | 3.7 (0.9) | 3.1 (0.9) |
| Pre-statin-treated[&] | 3.7 (0.9) | 4.0 (1.1) | 4.0 (0.9) | 4.1 (1.1) |
| HDL cholesterol (mmol/L) | 1.5 (0.4) | 1.4 (0.4) | 1.5 (0.4) | 1.3 (0.4) |
| Creatinine (umol/L) | 70.8 (13.9) | 73.4 (17.6) | 72.4 (16.7) | 78.7 (20.1) |
| Systolic BP (mmHg) | 133.6 (17.3) | 134.1 (17.7) | 144.2 (18.6) | 141.3 (19.1) |
| Diastolic BP (mmHg) | 82.2 (10.2) | 81.9 (10.5) | 82.8 (9.8) | 80.2 (10.5) |
| On antihypertensive treatment | 1 002 (10.0) | 3 276 (32.8) | 2 541 (25.4) | 5 225 (52.2) |
| History of diabetes | | | | |
| No | 9 625 (96.2) | 8 888 (88.9) | 9 348 (93.5) | 8 451 (84.5) |
| Type 1 | 52 (0.5) | 225 (2.2) | 55 (0.5) | 267 (2.7) |
| Type 2 | 323 (3.2) | 887 (8.9) | 597 (6.0) | 1 282 (12.8) |
| History of cancer | 508 (5.1) | 736 (7.4) | 1 049 (10.5) | 1 175 (11.8) |
| Severe mental illness history | 883 (8.8) | 1 457 (14.6) | 650 (6.5) | 929 (9.3) |
| History of CVD | | | | |

| | Age ≥40, <60 years | | Age ≥60, ≤70 years | |
|---|---|---|---|---|
| | **Without CVD history (n = 10 000)** | **With CVD history (n = 10 000)** | **Without CVD history (n = 10 000)** | **With CVD history (n = 10 000)** |
| No | 10 000 (100) | 0 (0) | 10 000 (100) | 0 (0) |
| MI only | 0 (0) | 325 (3.2) | 0 (0) | 393 (3.9) |
| Stroke only | 0 (0) | 978 (9.8) | 0 (0) | 818 (8.2) |
| PAD only | 0 (0) | 1 755 (17.5) | 0 (0) | 931 (9.3) |
| Other CHD only^ | 0 (0) | 4 949 (49.5) | 0 (0) | 5 046 (50.5) |
| Two or more | 0 (0) | 1 993 (19.9) | 0 (0) | 2 812 (28.1) |

BP, blood pressure; CHD, coronary heart disease; CVD, cardiovascular disease; HDL, high density lipoprotein; LDL, low density lipoprotein; PAD, peripheral arterial disease; SD, standard deviation. Values are mean (SD) or number (%).

*Other ethnicity includes Chinese, Mixed, White and Black Caribbean, White and Black African, White and Asian, Any other mixed background and other ethnic group.

^Other CHD includes acute rheumatic fever, chronic rheumatic heart diseases, hypertensive heart disease, angina pectoris, other acute ischaemic heart disease, chronic ischaemic heart disease, pulmonary heart disease and other form of heart disease.

&Pre-statin treated LDL cholesterol levels were calculated based on the baseline LDL cholesterol level, baseline statin usage and the LDL-reduction level by the statin.

*Supplementary table 3. Summary of adverse events and healthcare use during study follow-up*

| Parameter | Estimate | Source |
|---|---|---|
| **Relative reduction in LDL-C with atorvastatin 40 mg/day** | 49% | Law 2003[1] |
| **Effects of statin therapy on cardiovascular events per 1 mmol/L reduction in LDL-C, Rate Ratio (95% CI)** | | |
| Myocardial infarction | 0.76 (0.73-0.79) | CTT-C 2012[2] |
| Stroke | 0.84 (0.80-0.89) | |
| Coronary revascularization | 0.75 (0.73-0.78) | |
| Cardiovascular death | 0.88 (0.85-0.91) | |
| **Adverse effects of statin therapy** | | |
| Incident diabetes, Odds Ratios (95% CI) | | |
| Atovarstatin 20 mg/day vs. none | 1.09 (1.02-1.17) | Sattar 2010[3] |
| Atovarstatin 40 mg/day vs. 20 mg/day | 1.12 (1.04-1.22) | Preiss 2011[4] |
| Myopathy, excess rate per 100,000 treated with statin | 11 (4, 27) | Law 2006[5] |
| Rhabdomyolysis, excess rate per 100,000 treated with statin | 3.4 (1.6, 6.5) | |
| Rhabdomyolysis case fatality | 10% | |

***Supplementary figure 3. PSA convergence check upon key outcomes over lifetime per person***

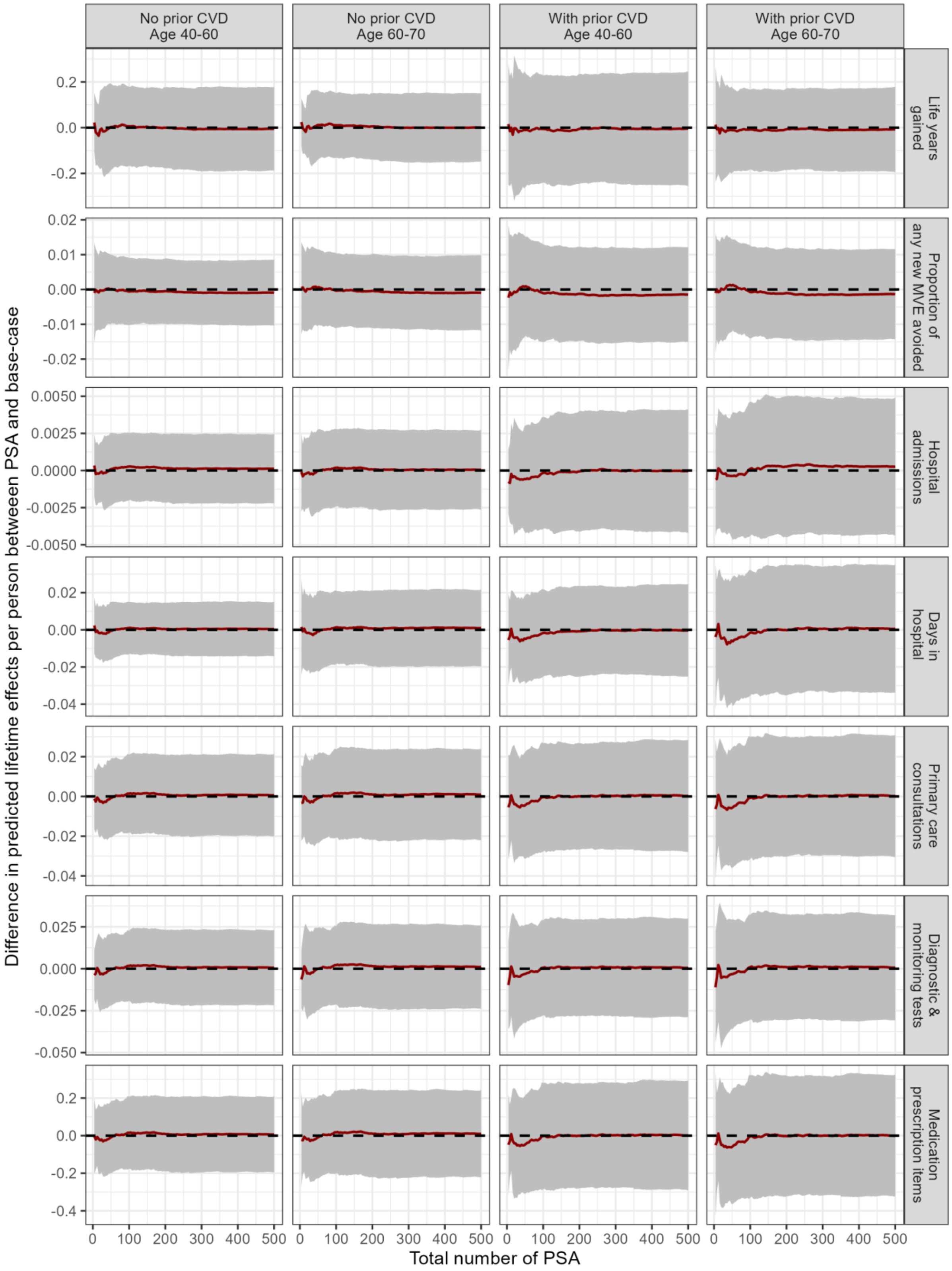

We ran 500 microsimulations per individual in the base-case and 100 microsimulations in the PSA. In total 500 PSAs were ran. For the outcome **per person**, PSA converges when the number of PSA is greater than 200. CVD = Cardiovascular disease, PSA = Probabilistic sensitivity analysis.

***Supplementary figure 4. PSA convergence check upon key outcomes over lifetime per person-year***

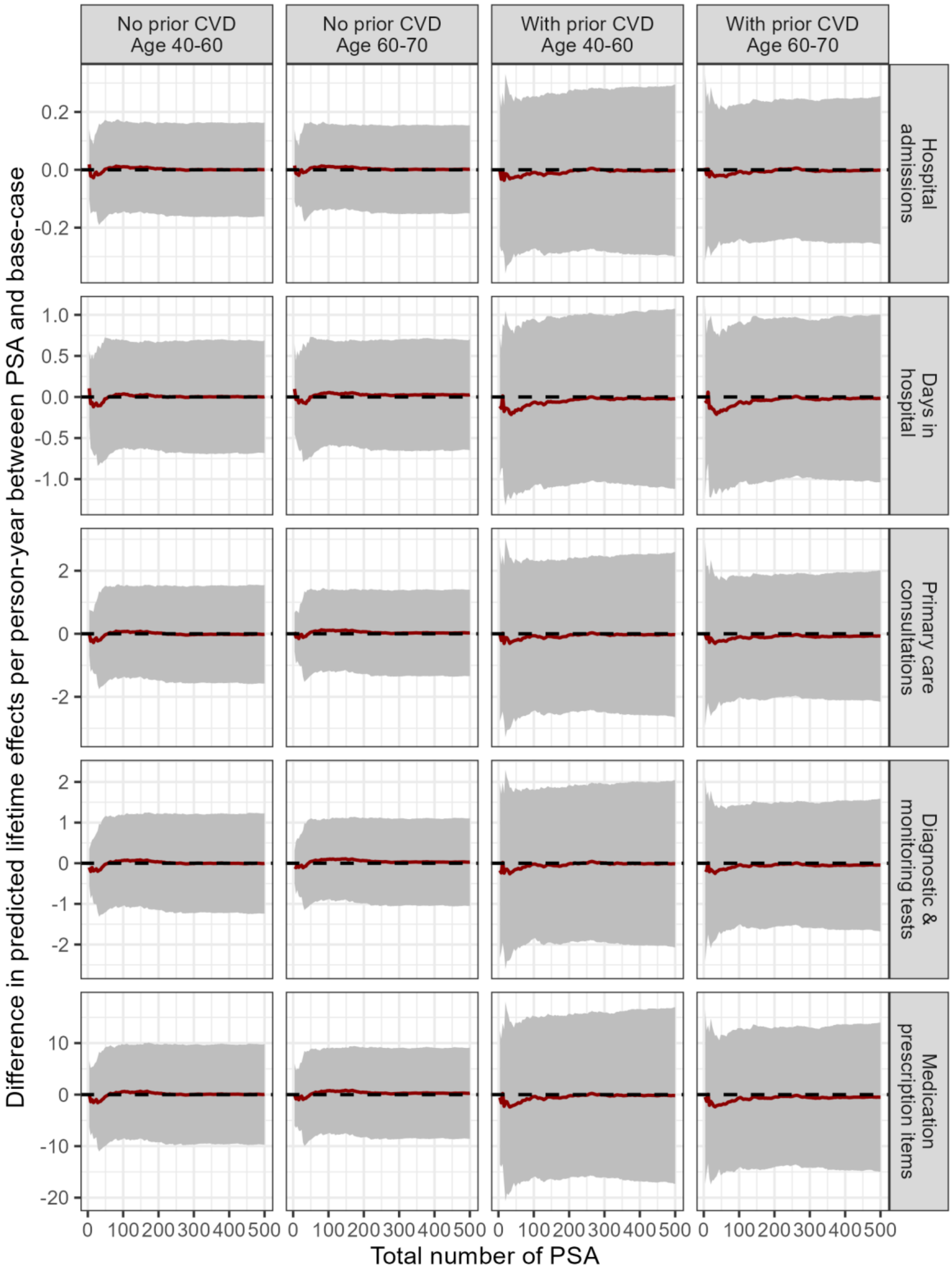

We ran 500 microsimulations per individual in the base-case and 100 microsimulations in the PSA. In total 500 PSAs were ran. For the outcome **per person-year**, PSA converges when the number of PSA is greater than 200. CVD = Cardiovascular disease, PSA = Probabilistic sensitivity analysis.

*Supplementary figure 5. A flowchart of inclusion of UK Biobank participants in annual primary and hospital care analyses*

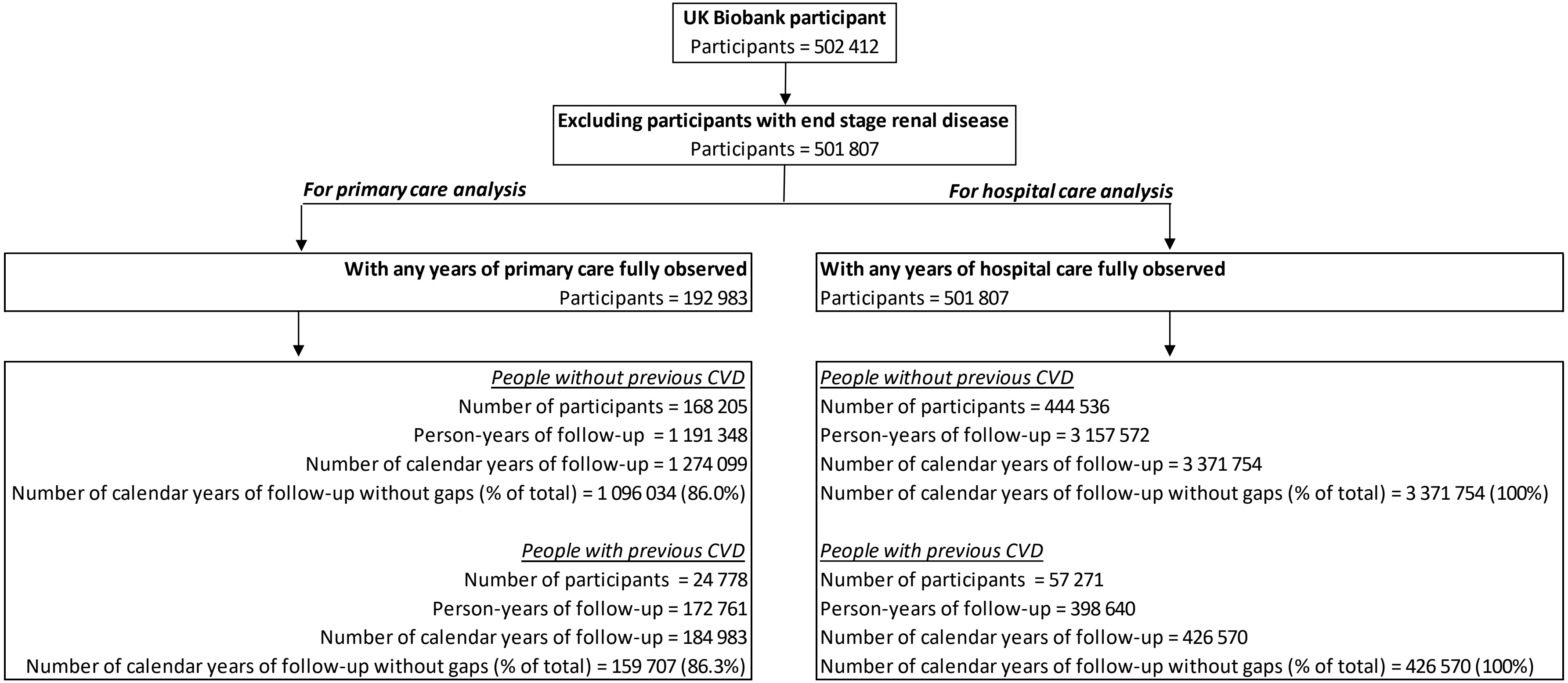

*Some participants may move to a primary care practice not contributing linked data to UK Biobank and would, therefore, have gaps in their registration records in the linked primary care data. Their annual periods with gaps in the primary care registrations were excluded from the primary care analysis.

***Supplementary table 4. Summary of adverse events and healthcare use during study follow-up***

| | **Primary care use analysis** | | **Hospital care use analysis** | |
|---|---|---|---|---|
| | **Without CVD history (n = 168 205)** | **With CVD history (n = 24 778)** | **Without CVD history (n = 444 536)** | **With history CVD (n = 57 271)** |
| Duration of follow-up (years) | 7.1 (0.9) | 7.0 (1.2) | 7.1 (1.0) | 7.0 (1.3) |
| *Number of participants with events during follow-up* | | | | |
| Myocardial infarction | 1 841 (1.1) | 871 (3.5) | 4 651 (1.0) | 2 210 (3.9) |
| Stroke | 1 730 (1.0) | 812 (3.3) | 4 106 (0.9) | 1 872 (3.3) |
| Coronary revascularization | 2 197 (1.3) | 1 205 (4.9) | 5 877 (1.3) | 3 094 (5.4) |
| Incident diabetes[&] | 3 425 (2.1) | 1 077 (5.0) | 7 395 (1.7) | 2 301 (4.7) |
| Incident cancer[&] | 9 057 (5.8) | 1 797 (8.0) | 25 376 (6.2) | 4 498 (8.7) |
| Vascular death | 521 (0.3) | 457 (1.8) | 1 781 (0.4) | 1 412 (2.5) |
| Non-vascular death | 2 777 (1.7) | 869 (3.5) | 9 067 (2.0) | 2 632 (4.6) |
| *Healthcare use during follow-up* | | | | |
| Total number of person-years used for analysis | 1 096 034 | 159 707 | 3 371 754 | 426 570 |
| Person- years with primary /hospital care use, respectively | 1 015 858 (92.7) | 156 379 (97.9) | 534 287 (15.8) | 121 167 (28.4) |
| Number of primary care consultations/ hospital inpatient episodes, respectively, per person-year (95% CIs) | 5.10 (5.08, 5.11) | 8.31 (8.24, 8.38) | 0.34 (0.33, 0.34) | 0.71 (0.70, 0.72) |
| Number of diagnostic and monitoring tests / days in hospital, respectively, per person-year (95% CIs) | 2.98 (2.96, 2.99) | 5.84 (5.77, 5.91) | 0.70 (0.70, 0.70) | 1.88 (1.87, 1.88) |
| Number of medication prescription items per person-year (95% CIs) | 17.9 (17.8, 18.0) | 48.3 (47.6, 49.0) | - | - |

CVD, cardiovascular disease; CI, confidence interval; SD, standard deviation.
Values are mean (SD) or number (%) unless stated otherwise.
[&]Calculated as proportion from participants without condition at baseline;

***Supplementary table 5. Two-part models of annual hospital care use associated with CVD events (Part 1: Logistic regression model; Part 2: Generalized linear model with Gamma distribution and identify link function)***

| **Covariate** | **Part 1: Likelihood of incurring any use Odds ratio (95%CIs) [Hospital admissions/Days in hospital]** | **Part 2: Number of use, if any incurred Mean (95%CIs)** | |
|---|---|---|---|
| | | **Hospital admissions** | **Days in hospital** |
| Intercept [a] | 0.13 (0.13, 0.14) | 1.5 (1.4, 1.6) | 2.4 (2.2, 2.5) |
| **BASELINE CHARACTERISTICS** | | | |
| Male (ref: female) | 0.91 (0.90, 0.92) | -0.1 (-0.1, 0.0) | 0.2 (0.1, 0.3) |
| Ethnicity (ref: white) | | | |
| Black | 1.04 (1.01, 1.08) | 0.0 (-0.1, 0.1) | 0.0 (-0.3, 0.3) |
| South Asian | 1.16 (1.12, 1.19) | 0.0 (-0.1, 0.1) | -0.2 (-0.4, 0.0) |
| Others* | 1.04 (1.01, 1.06) | 0.0 (-0.1, 0.0) | -0.2 (-0.3, 0.0) |
| Townsend socioeconomic deprivation (ref: Quintile 3) | | | |
| Quintile 1 (least deprived) | 0.94 (0.93, 0.96) | 0.0 (0.0, 0.0) | -0.2 (-0.3, -0.1) |
| Quintile 2 | 0.99 (0.97, 1.00) | 0.0 (0.0, 0.0) | -0.2 (-0.2, -0.1) |
| Quintile 4 | 1.06 (1.05, 1.08) | 0.0 (0.0, 0.0) | 0.2 (0.1, 0.3) |
| Quintile 5 | 1.16 (1.15, 1.18) | 0.1 (0.0, 0.1) | 0.6 (0.5, 0.8) |
| Smoking (ref: never) | | | |
| Former smoker | 1.10 (1.09, 1.11) | 0.0 (0.0, 0.0) | 0.0 (-0.1, 0.0) |
| Current smoker | 1.19 (1.18, 1.21) | 0.1 (0.0, 0.1) | 0.6 (0.4, 0.7) |
| Physical activity (ref: moderate) | | | |
| Low | 1.13 (1.11, 1.14) | 0.1 (0.0, 0.1) | 0.4 (0.3, 0.5) |
| High | 1.06 (1.05, 1.07) | 0.0 (0.0, 0.0) | -0.1 (-0.1, 0.0) |
| Missing | 1.15 (1.13, 1.16) | 0.0 (0.0, 0.0) | 0.2 (0.1, 0.3) |
| Unhealthy diet (ref: Healthy diet) | 1.06 (1.05, 1.07) | 0.0 (0.0, 0.0) | 0.1 (0.0, 0.1) |
| Body mass index (kg/m$^2$) (ref: ≥18.5, <25) | | | |
| <18.5 | 1.17 (1.10, 1.23) | 0.4 (0.0, 0.8) | 1.6 (0.8, 2.3) |
| ≥25, <30 | 1.11 (1.10, 1.12) | 0.0 (0.0, 0.0) | -0.1 (-0.2, 0.0) |
| ≥30, <35 | 1.23 (1.21, 1.24) | 0.0 (0.0, 0.0) | 0.1 (0.0, 0.2) |
| ≥35, <40 | 1.34 (1.31, 1.36) | 0.1 (0.0, 0.1) | 0.5 (0.3, 0.6) |
| ≥40 | 1.48 (1.44, 1.53) | 0.1 (0.0, 0.2) | 1.1 (0.6, 1.5) |

| Covariate | Part 1: Likelihood of incurring any use<br>Odds ratio (95%CIs)<br>[Hospital admissions/Days in hospital] | Part 2: Number of use, if any incurred<br>Mean (95%CIs) | |
|---|---|---|---|
| | | Hospital admissions | Days in hospital |
| LDL cholesterol (centred at 3.6; per 1 mmol/L) | 0.97 (0.97, 0.98) | 0.0 (0.0, 0.0) | -0.1 (-0.1, 0.0) |
| Natural logarithm of HDL cholesterol (lnmmol/L) | 0.89 (0.87, 0.91) | -0.1 (-0.1, 0.0) | -0.1 (-0.2, 0.0) |
| Natural logarithm of serum creatinine (centred at 4.4; per 0.2 lnumol/L) | 0.99 (0.99, 1.00) | 0.1 (0.0, 0.1) | 0.0 (-0.1, 0.1) |
| Systolic blood pressure (centred at 140; per 20 mmHg) | 0.94 (0.93, 0.95) | 0.0 (0.0, 0.0) | 0.0 (-0.1, 0.0) |
| Diastolic blood pressure (centred at 80; per 10 mmHg) | 1.01 (1.01, 1.02) | 0.0 (0.0, 0.0) | 0.0 (0.0, 0.0) |
| On antihypertensive treatment (ref: no) | 1.13 (1.12, 1.14) | 0.1 (0.0, 0.1) | 0.2 (0.1, 0.3) |
| Severe mental illness history (ref: no) | 1.79 (1.71, 1.87) | 0.5 (0.3, 0.8) | 2.1 (1.5, 2.6) |
| History of type 1 diabetes (ref: no) | 1.42 (1.40, 1.44) | 0.1 (0.1, 0.1) | 1.5 (1.3, 1.7) |
| History of CVD (ref: no) | | | |
| MI only | 1.17 (1.11, 1.24) | -0.1 (-0.1, 0.0) | 0.3 (-0.1, 0.7) |
| PAD only | 1.51 (1.46, 1.56) | 0.2 (0.1, 0.3) | 1.0 (0.7, 1.2) |
| Stroke only | 1.34 (1.29, 1.39) | 0.1 (0.0, 0.1) | 0.9 (0.6, 1.3) |
| Other CHD only^ | 1.53 (1.50, 1.55) | 0.1 (0.1, 0.1) | 0.3 (0.2, 0.4) |
| Two or more | 1.66 (1.62, 1.70) | 0.2 (0.1, 0.2) | 1.1 (0.8, 1.4) |
| **TIME-UPDATED CHARACTERISTICS** | | | |
| Current age (centred at 60; per 10 years) | 1.36 (1.35, 1.37) | 0.0 (0.0, 0.0) | 0.3 (0.3, 0.4) |
| Incident MI (ref: no) | | | |
| Same year | 47.9 (40.3, 56.9) | 0.4 (0.3, 0.5) | 7.2 (6.4, 8.0) |
| 1 year ago | 2.42 (2.17, 2.71) | 0.4 (0.3, 0.6) | 2.8 (1.7, 3.9) |
| ≥2 years ago | 1.65 (1.51, 1.82) | 0.2 (0.1, 0.4) | 2.4 (1.1, 3.7) |
| Incident Stroke (ref: no) | | | |
| Same year | 47.0 (42.4, 52.2) | 0.2 (0.1, 0.3) | 15.5 (14.6, 16.4) |
| 1 year ago | 2.43 (2.28, 2.60) | 0.2 (0.1, 0.4) | 10.4 (8.7, 12.2) |
| ≥2 years ago | 1.54 (1.45, 1.62) | 0.2 (0.0, 0.4) | 5.1 (3.6, 6.6) |
| Incident CRV (ref: no) | | | |
| Same year | -[b] | 0.5 (0.5, 0.6) | 3.5 (3.2, 3.7) |
| 1 year ago | 1.81 (1.69, 1.93) | 0.1 (0.0, 0.1) | 0.2 (-0.3, 0.7) |

| Covariate | Part 1: Likelihood of incurring any use Odds ratio (95%CIs) [Hospital admissions/Days in hospital] | Part 2: Number of use, if any incurred Mean (95%CIs) | |
|---|---|---|---|
| | | Hospital admissions | Days in hospital |
| ≥2 years ago | 1.42 (1.35, 1.50) | 0.0 (0.0, 0.1) | -0.1 (-0.4, 0.2) |
| Diabetes (ref: no) | | | |
| <10 years ago | 1.36 (1.34, 1.38) | 0.2 (0.1, 0.2) | 0.8 (0.6, 0.9) |
| ≥10 years ago | 1.20 (1.17, 1.23) | 0.1 (0.0, 0.2) | 0.8 (0.6, 1.1) |
| Cancer (ref: no) | | | |
| <5 years ago | 5.07 (5.00, 5.15) | 2.3 (2.3, 2.4) | 4.7 (4.5, 4.8) |
| ≥5 years ago | 1.68 (1.65, 1.70) | 0.8 (0.8, 0.9) | 1.6 (1.5, 1.7) |
| VD (ref = no) | 2.34 (2.12, 2.57) | 0.3 (0.1, 0.5) | 12.5 (10.6, 14.3) |
| NVD (ref = no) | 10.7 (10.1, 11.3) | 1.5 (1.4, 1.7) | 18.6 (18.0, 19.1) |
| **EVENT INTERACTIONS** | | | |
| *Interaction between temporal history of MI and CRV* | | | |
| MI: same year; CRV: same year (ref = no) | -[b] | -0.6 (-0.8, -0.5) | -5.4 (-6.3, -4.5) |
| MI: same year; CRV: 1 year ago (ref = no) | 0.38 (0.16, 0.91) | -0.1 (-0.6, 0.4) | -2.6 (-4.9, -0.2) |
| MI: same year; CRV: ≥2 years ago (ref = no) | 1.28 (0.56, 2.91) | 0.1 (-0.3, 0.4) | -3.1 (-5.2, -1.1) |
| MI: 1 year ago; CRV: same year (ref = no) | -[b] | -0.4 (-0.8, -0.1) | 0.2 (-2.0, 2.4) |
| MI: 1 year ago; CRV: 1 year ago (ref = no) | 0.56 (0.48, 0.65) | -0.2 (-0.4, 0.1) | -2.2 (-3.4, -0.9) |
| MI: 1 year ago; CRV: ≥2 years ago (ref = no) | 0.72 (0.49, 1.05) | -0.3 (-0.8, 0.1) | -3.2 (-5.0, -1.4) |
| MI: ≥2 years ago; CRV: same year (ref = no) | -[b] | 0.1 (-0.6, 0.9) | 1.8 (-2.0, 5.7) |
| MI: ≥2 years ago; CRV: 1 year ago (ref = no) | 0.82 (0.57, 1.18) | -0.1 (-0.6, 0.3) | -1.3 (-3.6, 1.1) |
| MI: ≥2 years ago; CRV: ≥2 years ago (ref = no) | 0.70 (0.62, 0.80) | -0.2 (-0.3, 0.0) | -2.1 (-3.5, -0.7) |
| *Interaction between CVD events and VD* | | | |
| Same year MI and same year VD (ref = no) | 0.02 (0.02, 0.03) | -0.6 (-1.0, -0.3) | -12.4 (-15.2, -9.6) |
| Same year CRV and same year VD (ref = no) | -[b] | -0.1 (-0.4, 0.3) | 5.4 (0.8, 10.1) |
| Same year Stroke and same year VD (ref = no) | 0.16 (0.12, 0.22) | -0.6 (-0.8, -0.4) | -15.6 (-18.5, -12.7) |

CHD, coronary heart disease; CRV, coronary revascularization; CVD, cardiovascular disease; HDL, high density lipoprotein; LDL, low density lipoprotein; MI, myocardial infarction; NVD, non-vascular death; PAD, peripheral arterial disease; VD, vascular death;

Resource use in years with administrative censoring at end of study follow-up were adjusted by including a further covariate of proportion of year observed (not shown).

* Other ethnicity includes Chinese, Mixed, White and Black Caribbean, White and Black African, White and Asian, Any other mixed background and other ethnic group;

^ Other CHD includes acute rheumatic fever, chronic rheumatic heart diseases, hypertensive heart disease, angina pectoris, other acute ischaemic heart disease, chronic ischaemic heart disease, pulmonary heart disease and other form of heart disease

[a] The intercept terms represent the mean odds of any none-zero use [=probability/(1-probability)] (or mean number of use conditional on any none-zero use) for an individual in the reference categories of covariates; Other coefficients represent the odds ratio (or mean difference) for that category of the covariate compared to the reference category.

[b] People with same year CRV always have none-zero number of hospital admission or days in hospital.

***Supplementary table 6. Two-part models of annual primary care use associated with CVD events (Part 1: Logistic regression model; Part 2: Generalized linear model with Gamma distribution and identify link function)***

| Covariates | Primary care consultations | | Diagnostic and monitoring tests | | Medication prescription items | |
|---|---|---|---|---|---|---|
| | **Part 1 OR (95%CIs)** | **Part 2 Mean (95%CIs)** | **Part 1 OR (95%CIs)** | **Part 2 Mean (95%CIs)** | **Part 1 OR (95%CIs)** | **Part 2 Mean (95%CIs)** |
| Intercept [a] | 11.4 (11.0, 11.9) | 4.7 (4.6, 4.7) | 2.50 (2.43, 2.57) | 3.8 (3.7, 3.9) | 3.03 (2.91, 3.15) | 13.3 (12.8, 13.8) |
| **BASELINE CHARACTERISTICS** | | | | | | |
| Male (ref: female) | 0.53 (0.52, 0.55) | -0.5 (-0.5, -0.5) | 0.48 (0.47, 0.49) | -0.3 (-0.3, -0.3) | 0.65 (0.64, 0.67) | -1.2 (-1.5, -0.9) |
| Ethnicity (ref: white) | | | | | | |
| Black | 1.31 (1.19, 1.43) | 0.5 (0.4, 0.7) | 1.31 (1.23, 1.39) | 0.5 (0.3, 0.6) | 1.51 (1.39, 1.65) | -1.3 (-2.2, -0.5) |
| South Asian | 1.62 (1.50, 1.75) | 0.8 (0.7, 1.0) | 1.81 (1.72, 1.91) | 0.7 (0.6, 0.8) | 2.09 (1.94, 2.25) | 1.5 (0.9, 2.2) |
| Others* | 1.14 (1.07, 1.22) | 0.2 (0.1, 0.3) | 1.22 (1.16, 1.27) | 0.3 (0.3, 0.4) | 1.22 (1.15, 1.30) | -0.7 (-1.4, 0.0) |
| Townsend socioeconomic deprivation (ref: Quintile 3) | | | | | | |
| Quintile 1 (least deprived) | 1.01 (0.98, 1.03) | -0.1 (-0.2, -0.1) | 0.99 (0.97, 1.01) | -0.1 (-0.1, 0.0) | 1.00 (0.98, 1.03) | -1.4 (-1.8, -1.0) |
| Quintile 2 | 1.01 (0.98, 1.04) | -0.1 (-0.2, -0.1) | 1.00 (0.98, 1.02) | 0.0 (-0.1, 0.0) | 1.03 (1.00, 1.06) | -1.1 (-1.5, -0.6) |
| Quintile 4 | 1.04 (1.01, 1.08) | 0.0 (0.0, 0.0) | 1.03 (1.01, 1.05) | 0.1 (0.0, 0.1) | 0.99 (0.96, 1.02) | -0.1 (-0.5, 0.3) |
| Quintile 5 | 1.10 (1.06, 1.14) | 0.3 (0.2, 0.3) | 1.03 (1.01, 1.06) | 0.1 (0.1, 0.2) | 1.05 (1.01, 1.09) | 2.7 (2.2, 3.2) |
| Smoking (ref: never) | | | | | | |
| Former smoker | 1.21 (1.19, 1.24) | 0.2 (0.2, 0.2) | 1.13 (1.11, 1.14) | 0.1 (0.1, 0.2) | 1.20 (1.18, 1.22) | 0.8 (0.6, 1.0) |
| Current smoker | 1.23 (1.20, 1.27) | 0.4 (0.3, 0.4) | 1.06 (1.04, 1.08) | 0.2 (0.1, 0.2) | 1.19 (1.16, 1.23) | 2.4 (2.0, 2.7) |
| Physical activity (ref: moderate) | | | | | | |
| Low | 1.06 (1.03, 1.09) | 0.2 (0.2, 0.3) | 1.06 (1.04, 1.08) | 0.2 (0.2, 0.2) | 1.12 (1.09, 1.15) | 1.6 (1.3, 1.9) |
| High | 1.04 (1.02, 1.06) | 0.0 (0.0, 0.1) | 1.01 (0.99, 1.02) | 0.0 (0.0, 0.0) | 1.03 (1.01, 1.05) | 0.0 (-0.3, 0.3) |
| Missing | 1.11 (1.08, 1.14) | 0.3 (0.2, 0.3) | 1.07 (1.05, 1.09) | 0.2 (0.1, 0.2) | 1.14 (1.11, 1.17) | 1.8 (1.5, 2.1) |
| Unhealthy diet (ref: Healthy diet) | 1.02 (1.00, 1.04) | 0.1 (0.0, 0.1) | 0.99 (0.98, 1.00) | 0.1 (0.0, 0.1) | 1.01 (0.99, 1.03) | 0.3 (0.1, 0.5) |
| Body mass index (kg/m$^2$) (ref: ≥18.5, <25) | | | | | | |
| <18.5 | 1.03 (0.91, 1.16) | 0.4 (0.2, 0.6) | 1.06 (0.98, 1.16) | 0.4 (0.2, 0.7) | 1.05 (0.93, 1.18) | 2.0 (0.9, 3.1) |
| ≥25, <30 | 1.09 (1.07, 1.11) | 0.1 (0.1, 0.1) | 1.08 (1.06, 1.09) | 0.1 (0.0, 0.1) | 1.14 (1.12, 1.17) | 0.7 (0.5, 1.0) |
| ≥30, <35 | 1.19 (1.15, 1.22) | 0.5 (0.4, 0.5) | 1.17 (1.15, 1.19) | 0.3 (0.2, 0.3) | 1.29 (1.25, 1.32) | 3.2 (2.8, 3.5) |
| ≥35, <40 | 1.22 (1.16, 1.28) | 1.0 (0.9, 1.1) | 1.24 (1.20, 1.28) | 0.5 (0.4, 0.6) | 1.34 (1.28, 1.41) | 6.4 (5.8, 7.0) |
| ≥40 | 1.41 (1.28, 1.55) | 1.6 (1.5, 1.8) | 1.33 (1.26, 1.41) | 0.7 (0.6, 0.9) | 1.43 (1.31, 1.56) | 11.8 (10.6, 13.1) |

| Covariates | Primary care consultations | | Diagnostic and monitoring tests | | Medication prescription items | |
|---|---|---|---|---|---|---|
| | **Part 1 OR (95%CIs)** | **Part 2 Mean (95%CIs)** | **Part 1 OR (95%CIs)** | **Part 2 Mean (95%CIs)** | **Part 1 OR (95%CIs)** | **Part 2 Mean (95%CIs)** |
| LDL cholesterol (centred at 3.6; per 1 mmol/L) | 0.94 (0.93, 0.95) | -0.1 (-0.1, 0.0) | 0.92 (0.91, 0.93) | 0.0 (-0.1, 0.0) | 0.93 (0.92, 0.94) | -0.4 (-0.6, -0.3) |
| Natural logarithm of HDL cholesterol (lnmmol/L) | 0.75 (0.72, 0.78) | -0.4 (-0.5, -0.3) | 0.75 (0.73, 0.77) | -0.5 (-0.5, -0.4) | 0.69 (0.66, 0.72) | -1.7 (-2.3, -1.1) |
| Natural logarithm of serum creatinine (centred at 4.4; per 0.2 lnumol/L) | 0.98 (0.97, 1.00) | 0.0 (0.0, 0.0) | 1.03 (1.02, 1.04) | 0.1 (0.1, 0.1) | 1.00 (0.98, 1.01) | -0.3 (-0.5, -0.2) |
| Systolic blood pressure (centred at 140; per 20 mmHg) | 0.96 (0.95, 0.97) | 0.1 (0.1, 0.1) | 0.98 (0.97, 0.99) | 0.0 (0.0, 0.0) | 0.95 (0.94, 0.96) | 0.3 (0.1, 0.4) |
| Diastolic blood pressure (centred at 80; per 10 mmHg) | 1.07 (1.05, 1.08) | 0.0 (0.0, 0.0) | 1.06 (1.05, 1.07) | 0.0 (0.0, 0.0) | 1.07 (1.06, 1.09) | 0.2 (0.0, 0.4) |
| On antihypertensive treatment (ref: no) | 2.93 (2.83, 3.03) | 1.3 (1.2, 1.3) | 2.88 (2.82, 2.94) | 0.4 (0.4, 0.4) | 3.12 (3.01, 3.23) | 15.8 (15.4, 16.1) |
| Severe mental illness history (ref: no) | 2.21 (1.67, 2.92) | 1.8 (1.5, 2.1) | 1.72 (1.48, 2.01) | 0.6 (0.3, 0.8) | 1.35 (1.08, 1.68) | 19.9 (17.0, 22.7) |
| History of type 1 diabetes (ref: no) | 1.96 (1.89, 2.03) | 1.3 (1.2, 1.4) | 1.45 (1.42, 1.48) | 0.5 (0.5, 0.6) | 2.03 (1.96, 2.10) | 9.5 (9.0, 9.9) |
| History of CVD (ref: no) | | | | | | |
| MI only | 1.76 (1.45, 2.15) | 0.7 (0.5, 1.0) | 2.34 (2.07, 2.65) | 0.2 (-0.1, 0.4) | 2.04 (1.64, 2.53) | 12.0 (10.2, 13.8) |
| PAD only | 1.51 (1.40, 1.63) | 1.0 (0.8, 1.1) | 1.36 (1.30, 1.43) | 0.7 (0.6, 0.9) | 1.46 (1.36, 1.57) | 6.4 (5.5, 7.3) |
| Stroke only | 1.77 (1.55, 2.02) | 1.2 (1.0, 1.3) | 1.95 (1.80, 2.10) | 0.5 (0.3, 0.6) | 1.58 (1.39, 1.78) | 14.4 (12.8, 15.9) |
| Other CHD only^ | 1.60 (1.52, 1.67) | 1.5 (1.4, 1.5) | 1.73 (1.68, 1.78) | 1.6 (1.5, 1.7) | 1.57 (1.50, 1.64) | 11.1 (10.5, 11.6) |
| Two or more | 2.07 (1.89, 2.27) | 2.1 (2.0, 2.3) | 2.86 (2.71, 3.03) | 2.0 (1.9, 2.2) | 1.74 (1.59, 1.91) | 30.7 (29.4, 32.0) |
| **TIME-UPDATED CHARACTERISTICS** | | | | | | |
| Current age (centred at 60; per 10 years) | 1.98 (1.96, 2.01) | 0.6 (0.6, 0.6) | 1.46 (1.45, 1.47) | 0.4 (0.4, 0.5) | 1.67 (1.65, 1.69) | 3.8 (3.7, 3.9) |
| Incident MI (ref: no) | | | | | | |
| Same year | 4.87 (2.51, 9.45) | 5.0 (4.5, 5.6) | 3.95 (2.89, 5.40) | 2.7 (2.2, 3.2) | 1.86 (1.39, 2.50) | 21.4 (18.5, 24.3) |
| 1 year ago | 2.67 (1.65, 4.31) | 3.1 (2.6, 3.7) | 3.41 (2.51, 4.62) | 2.3 (1.7, 2.9) | 2.18 (1.54, 3.07) | 35.9 (31.6, 40.1) |
| ≥2 years ago | 1.53 (1.14, 2.04) | 1.3 (0.9, 1.8) | 2.09 (1.70, 2.56) | 1.5 (0.9, 2.0) | 2.38 (1.70, 3.33) | 29.8 (24.6, 34.9) |
| Incident Stroke (ref: no) | | | | | | |
| Same year | 12.1 (6.64, 22.0) | 4.8 (4.5, 5.1) | 5.77 (4.70, 7.09) | 2.7 (2.4, 3.0) | 2.22 (1.85, 2.66) | 15.3 (13.5, 17.2) |
| 1 year ago | 2.66 (2.00, 3.55) | 2.7 (2.4, 3.0) | 2.95 (2.51, 3.47) | 2.1 (1.7, 2.4) | 2.19 (1.80, 2.65) | 25.5 (22.9, 28.1) |
| ≥2 years ago | 1.56 (1.33, 1.83) | 1.5 (1.2, 1.8) | 2.09 (1.86, 2.35) | 1.8 (1.4, 2.1) | 2.43 (2.04, 2.90) | 23.5 (19.5, 27.5) |
| Incident CRV (ref: no) | | | | | | |
| Same year | 40.2 (10.0, 162.1) | 6.3 (5.9, 6.6) | 13.4 (9.33, 19.3) | 2.7 (2.4, 3.0) | 2.55 (2.01, 3.23) | 27.0 (25.1, 28.8) |
| 1 year ago | 3.60 (2.44, 5.30) | 2.3 (2.0, 2.6) | 4.94 (3.89, 6.28) | 1.2 (0.9, 1.5) | 2.89 (2.23, 3.74) | 28.5 (26.6, 30.5) |
| ≥2 years ago | 1.75 (1.42, 2.16) | 0.8 (0.6, 1.0) | 2.71 (2.37, 3.11) | 0.7 (0.4, 0.9) | 3.58 (2.81, 4.56) | 21.0 (19.0, 23.1) |

| Covariates | Primary care consultations | | Diagnostic and monitoring tests | | Medication prescription items | |
|---|---|---|---|---|---|---|
| | Part 1 OR (95%CIs) | Part 2 Mean (95%CIs) | Part 1 OR (95%CIs) | Part 2 Mean (95%CIs) | Part 1 OR (95%CIs) | Part 2 Mean (95%CIs) |
| Diabetes (ref: no) | | | | | | |
| <10 years ago | 2.74 (2.52, 2.97) | 2.9 (2.8, 2.9) | 4.43 (4.21, 4.67) | 3.0 (2.9, 3.0) | 2.07 (1.94, 2.22) | 20.6 (19.8, 21.5) |
| ≥10 years ago | 2.08 (1.88, 2.31) | 2.8 (2.6, 2.9) | 3.97 (3.71, 4.25) | 2.9 (2.8, 3.0) | 2.12 (1.90, 2.36) | 31.6 (30.3, 32.9) |
| Cancer (ref: no) | | | | | | |
| <5 years ago | 2.27 (2.14, 2.41) | 2.2 (2.1, 2.3) | 2.01 (1.94, 2.08) | 1.1 (1.0, 1.2) | 1.85 (1.76, 1.95) | 5.4 (5.0, 5.9) |
| ≥5 years ago | 1.29 (1.24, 1.34) | 0.6 (0.5, 0.6) | 1.25 (1.22, 1.28) | 0.3 (0.3, 0.4) | 1.32 (1.28, 1.37) | 3.1 (2.7, 3.5) |
| VD (ref = no) | 0.27 (0.20, 0.37) | -0.9 (-1.5, -0.4) | 0.33 (0.26, 0.42) | 0.2 (-0.4, 0.9) | 0.58 (0.45, 0.76) | -3.4 (-5.9, -0.9) |
| NVD (ref = no) | 0.36 (0.31, 0.42) | 1.8 (1.5, 2.1) | 0.41 (0.37, 0.46) | 0.8 (0.5, 1.0) | 0.72 (0.64, 0.81) | 13.4 (11.3, 15.4) |
| **EVENT INTERACTIONS** | | | | | | |
| *Interaction between temporal history of MI and CRV* | | | | | | |
| MI: same year; CRV: same year (ref = no) | 0.12 (0.02, 0.70) | -5.5 (-6.2, -4.8) | 0.22 (0.13, 0.40) | -2.9 (-3.6, -2.3) | 0.68 (0.44, 1.06) | -20.9 (-25.1, -16.8) |
| MI: same year; CRV: 1 year ago (ref = no) | 191 (63, 580) | -2.7 (-5.9, 0.5) | 123 (51, 294) | -3.8 (-5.8, -1.8) | 0.15 (0.02, 1.02) | -11.4 (-29.7, 6.9) |
| MI: same year; CRV: ≥2 years ago (ref = no) | 0.24 (0.02, 2.38) | -2.2 (-4.2, -0.2) | 0.19 (0.05, 0.81) | -2.2 (-4.3, -0.2) | 0.10 (0.03, 0.39) | -15.5 (-28.7, -2.4) |
| MI: 1 year ago; CRV: same year (ref = no) | 55.6 (11.8, 262.0) | -2.7 (-4.6, -0.7) | 0.15 (0.04, 0.56) | -2.4 (-3.9, -0.9) | 0.88 (0.19, 4.13) | -22.5 (-32.5, -12.6) |
| MI: 1 year ago; CRV: 1 year ago (ref = no) | 0.72 (0.34, 1.53) | -2.6 (-3.4, -1.9) | 0.41 (0.26, 0.64) | -1.9 (-2.6, -1.1) | 0.58 (0.35, 0.95) | -24.3 (-29.8, -18.9) |
| MI: 1 year ago; CRV: ≥2 years ago (ref = no) | 0.44 (0.06, 3.18) | -2.7 (-4.4, -1.1) | 0.29 (0.07, 1.26) | -3.1 (-4.6, -1.6) | 0.16 (0.04, 0.68) | -28.3 (-42.6, -14.1) |
| MI: ≥2 years ago; CRV: same year (ref = no) | 48.6 (9.6, 247.1) | -0.9 (-9.3, 7.5) | 0.08 (0.01, 0.69) | -0.4 (-8.7, 8.0) | 73.6 (30.1, 180.2) | -22.0 (-57.3, 13.2) |
| MI: ≥2 years ago; CRV: 1 year ago (ref = no) | 0.18 (0.06, 0.53) | 0.1 (-2.3, 2.5) | 0.16 (0.06, 0.40) | -0.6 (-2.7, 1.5) | 0.31 (0.10, 0.98) | -18.9 (-29.9, -7.9) |
| MI: ≥2 years ago; CRV: ≥2 years ago (ref = no) | 0.84 (0.56, 1.24) | -0.9 (-1.5, -0.4) | 0.55 (0.42, 0.72) | -1.4 (-2.0, -0.7) | 0.60 (0.37, 0.96) | -23.4 (-29.4, -17.3) |
| *Interaction between CVD events and VD* | | | | | | |
| Same year MI and same year VD (ref = no) | 0.13 (0.06, 0.32) | -5.8 (-6.7, -4.8) | 0.22 (0.13, 0.39) | -2.8 (-4.2, -1.5) | 0.36 (0.20, 0.65) | -23.3 (-28.9, -17.8) |
| Same year CRV and same year VD (ref = no) | 0.87 (0.12, 6.11) | -0.4 (-2.1, 1.3) | 0.72 (0.18, 2.85) | 0.5 (-1.5, 2.5) | 0.66 (0.21, 2.10) | -3.7 (-12.9, 5.5) |
| Same year Stroke and same year VD (ref = no) | 0.07 (0.03, 0.16) | -4.6 (-5.7, -3.5) | 0.15 (0.09, 0.25) | -2.9 (-4.1, -1.7) | 0.50 (0.28, 0.88) | -17.4 (-21.3, -13.4) |

CHD, coronary heart disease; CRV, coronary revascularization; CVD, cardiovascular disease; HDL, high density lipoprotein; LDL, low density lipoprotein; MI, myocardial infarction; NVD, non-vascular death; OR, odd ratio; PAD, peripheral arterial disease; VD, vascular death;

Resource use in years with administrative censoring at end of study follow-up were adjusted by including a further covariate of proportion of year observed (not shown).

* Other ethnicity includes Chinese, Mixed, White and Black Caribbean, White and Black African, White and Asian, Any other mixed background and other ethnic group;

^ Other CHD includes acute rheumatic fever, chronic rheumatic heart diseases, hypertensive heart disease, angina pectoris, other acute ischaemic heart disease, chronic ischaemic heart disease, pulmonary heart disease and other form of heart disease

[a] The intercept terms represent the mean odds of any none-zero use [=probability/(1-probability)] (or mean number of use conditional on any none-zero use) for an individual in the reference categories of covariates; Other coefficients represent the odds ratio (or mean difference) for that category of the covariate compared to the reference category.

***Supplementary table 7. Temporal impact of cardiovascular events on healthcare resource use***

(a) absolute effect

| **Temporal history of cardiovascular events (ref: none)** | **Extra annual healthcare resource use, mean (95% CIs)** | | | | |
|---|---|---|---|---|---|
| | **Hospital admissions** | **Days in hospital** | **Primary care consultations** | **Diagnostic and monitoring tests** | **Medication prescription items** |
| **MI** | | | | | |
| Same year | 1.4 (1.3, 1.5) | 8.5 (7.8, 9.2) | 5.2 (4.6, 5.7) | 3.1 (2.6, 3.5) | 19.5 (16.8, 22.2) |
| 1 year ago | 0.4 (0.3, 0.5) | 1.4 (1.0, 1.8) | 3.2 (2.6, 3.7) | 2.6 (2.1, 3.1) | 32.6 (28.5, 36.8) |
| ≥2 year ago | 0.2 (0.1, 0.2) | 0.9 (0.5, 1.2) | 1.3 (0.9, 1.7) | 1.6 (1.1, 2.1) | 27.7 (23.0, 32.4) |
| **Stroke** | | | | | |
| Same year | 1.3 (1.2, 1.3) | 15.8 (15.0, 16.7) | 5.2 (4.8, 5.5) | 3.3 (3.0, 3.5) | 15.1 (13.4, 16.8) |
| 1 year ago | 0.3 (0.3, 0.4) | 3.8 (3.2, 4.4) | 2.8 (2.5, 3.1) | 2.4 (2.1, 2.7) | 24.0 (21.6, 26.3) |
| ≥2 year ago | 0.1 (0.1, 0.2) | 1.4 (1.1, 1.8) | 1.6 (1.3, 1.8) | 1.9 (1.6, 2.2) | 22.6 (18.9, 26.4) |
| **CRV** | | | | | |
| Same year | 1.8 (1.8, 1.8) | 6.0 (5.7, 6.3) | 6.6 (6.3, 7.0) | 3.5 (3.2, 3.8) | 25.7 (23.8, 27.5) |
| 1 year ago | 0.2 (0.1, 0.2) | 0.4 (0.2, 0.5) | 2.5 (2.2, 2.8) | 1.8 (1.6, 2.1) | 27.5 (25.6, 29.4) |
| ≥2 year ago | 0.1 (0.1, 0.1) | 0.1 (0.1, 0.2) | 0.9 (0.7, 1.1) | 1.1 (0.9, 1.3) | 21.3 (19.3, 23.3) |
| **VD** | 0.3 (0.3, 0.4) | 4.3 (3.6, 5.0) | -1.4 (-1.9, -1.0) | -0.8 (-1.1, -0.4) | -4.0 (-5.9, -2.2) |

CRV: coronary re-vascularization; MI: myocardial infarction; VD: vascular death

(b) relative effect

| **Temporal history of cardiovascular events (ref: none)** | **Extra annual healthcare resource use, ratio (95% CIs)** | | | | |
|---|---|---|---|---|---|
| | **Hospital admissions** | **Days in hospital** | **Primary care consultations** | **Diagnostic and monitoring tests** | **Medication prescription items** |
| **MI** | | | | | |
| Same year | 5.50 (5.20, 5.79) | 12.28 (11.38, 13.19) | 2.03 (1.92, 2.14) | 1.99 (1.84, 2.14) | 2.00 (1.86, 2.14) |
| 1 year ago | 2.19 (1.96, 2.42) | 2.85 (2.36, 3.33) | 1.63 (1.52, 1.74) | 1.84 (1.67, 2.01) | 2.67 (2.46, 2.88) |
| ≥2 year ago | 1.56 (1.43, 1.70) | 2.14 (1.70, 2.58) | 1.26 (1.18, 1.35) | 1.53 (1.37, 1.68) | 2.42 (2.18, 2.66) |
| **Stroke** | | | | | |
| Same year | 4.99 (4.82, 5.16) | 22.75 (21.61, 23.89) | 2.02 (1.96, 2.08) | 2.06 (1.97, 2.15) | 1.78 (1.69, 1.86) |
| 1 year ago | 2.01 (1.84, 2.18) | 6.28 (5.47, 7.09) | 1.55 (1.49, 1.61) | 1.77 (1.67, 1.87) | 2.23 (2.11, 2.35) |
| ≥2 year ago | 1.47 (1.34, 1.60) | 2.95 (2.46, 3.43) | 1.31 (1.25, 1.36) | 1.61 (1.51, 1.71) | 2.16 (1.97, 2.36) |
| **CRV** | | | | | |
| Same year | 6.68 (6.55, 6.80) | 8.91 (8.57, 9.26) | 2.31 (2.25, 2.38) | 2.15 (2.05, 2.25) | 2.32 (2.23, 2.42) |
| 1 year ago | 1.52 (1.43, 1.61) | 1.48 (1.31, 1.66) | 1.50 (1.44, 1.56) | 1.59 (1.51, 1.68) | 2.41 (2.32, 2.51) |
| ≥2 year ago | 1.29 (1.22, 1.36) | 1.18 (1.10, 1.27) | 1.18 (1.13, 1.22) | 1.36 (1.29, 1.42) | 2.10 (1.99, 2.20) |
| **VD** | 2.04 (1.85, 2.24) | 7.10 (6.18, 8.03) | 0.72 (0.63, 0.80) | 0.75 (0.63, 0.87) | 0.79 (0.70, 0.89) |

CRV: coronary re-vascularization; MI: myocardial infarction; VD: vascular death

***Supplementary table 8. Predicted CVD events, life years and healthcare use over years in the scenario without statin treatment***

| Timeframe (years) | Clinical outcomes | | Healthcare use per person | | | | | Healthcare use per person year | | | | |
|---|---|---|---|---|---|---|---|---|---|---|---|---|
| | **% with new MVE** | **Life years per person** | **Hospital admissions** | **Hospital days** | **Consultations** | **Tests** | **Prescription items** | **Hospital admissions** | **Hospital days** | **Consultations** | **Tests** | **Prescription items** |
| **Without CVD History: Age 40-60 years** | | | | | | | | | | | | |
| 10 | 3.3% (3.2%, 3.4%) | 9.9 (9.9, 9.9) | 2.9 (2.9, 2.9) | 6.5 (6.4, 6.5) | 46 (46, 46) | 27 (27, 27) | 145 (144, 146) | 0.29 (0.29, 0.30) | 0.65 (0.65, 0.66) | 4.66 (4.64, 4.67) | 2.76 (2.75, 2.77) | 14.7 (14.6, 14.8) |
| 20 | 8.8% (8.5%, 9.1%) | 19.2 (19.1, 19.2) | 6.9 (6.8, 7.0) | 16.7 (16.5, 17.0) | 99 (98, 99) | 61 (60, 61) | 343 (341, 346) | 0.36 (0.36, 0.36) | 0.87 (0.86, 0.88) | 5.14 (5.12, 5.16) | 3.16 (3.14, 3.17) | 17.9 (17.8, 18.0) |
| 30 | 16.5% (15.4%, 17.6%) | 27.0 (26.8, 27.2) | 11.5 (11.3, 11.7) | 30.8 (30.2, 31.4) | 150 (148, 151) | 95 (94, 96) | 560 (552, 568) | 0.43 (0.42, 0.43) | 1.14 (1.12, 1.17) | 5.55 (5.52, 5.57) | 3.51 (3.48, 3.53) | 20.8 (20.6, 21.0) |
| Lifetime | 35.6% (29.7%, 41.6%) | 35.2 (33.9, 36.5) | 18.1 (17.0, 19.3) | 56.1 (52.1, 60.1) | 212 (202, 223) | 139 (131, 147) | 853 (797, 909) | 0.52 (0.50, 0.53) | 1.59 (1.52, 1.66) | 6.04 (5.94, 6.13) | 3.94 (3.86, 4.02) | 24.2 (23.5, 25.0) |
| **Without CVD History: Age 60-70 years** | | | | | | | | | | | | |
| 10 | 6.9% (6.8%, 7.1%) | 9.6 (9.6, 9.7) | 4.7 (4.7, 4.8) | 11.8 (11.7, 11.9) | 59 (59, 59) | 37 (37, 37) | 233 (231, 235) | 0.49 (0.49, 0.50) | 1.22 (1.21, 1.24) | 6.12 (6.10, 6.14) | 3.85 (3.83, 3.87) | 24.1 (24.0, 24.3) |
| 20 | 16.9% (16.3%, 17.4%) | 17.8 (17.7, 17.9) | 10.5 (10.4, 10.6) | 28.7 (28.2, 29.2) | 117 (117, 118) | 76 (76, 77) | 496 (491, 501) | 0.59 (0.58, 0.60) | 1.61 (1.58, 1.64) | 6.59 (6.56, 6.62) | 4.28 (4.25, 4.31) | 27.8 (27.6, 28.1) |
| 30 | 27.3% (25.7%, 28.9%) | 23.2 (22.9, 23.5) | 15.3 (15.0, 15.7) | 45.3 (44.3, 46.4) | 161 (158, 164) | 106 (105, 108) | 709 (694, 724) | 0.66 (0.65, 0.67) | 1.95 (1.91, 2.00) | 6.93 (6.88, 6.97) | 4.59 (4.55, 4.63) | 30.5 (30.2, 30.9) |
| Lifetime | 38.2% (34.6%, 41.7%) | 26.3 (25.5, 27.1) | 18.4 (17.6, 19.2) | 58.0 (55.1, 60.9) | 188 (181, 195) | 126 (121, 131) | 849 (809, 889) | 0.70 (0.68, 0.72) | 2.21 (2.14, 2.28) | 7.16 (7.08, 7.24) | 4.80 (4.73, 4.86) | 32.3 (31.6, 33.0) |
| **With CVD History: Age 40-60 years** | | | | | | | | | | | | |
| 10 | 13.8% (13.4%, 14.3%) | 9.6 (9.6, 9.7) | 5.6 (5.5, 5.7) | 15.5 (15.1, 15.8) | 71 (71, 72) | 50 (50, 51) | 378 (374, 383) | 0.58 (0.57, 0.59) | 1.61 (1.57, 1.64) | 7.42 (7.37, 7.47) | 5.24 (5.17, 5.30) | 39.2 (38.8, 39.7) |
| 20 | 27.1% (26.0%, 28.2%) | 17.9 (17.8, 18.0) | 12.0 (11.8, 12.2) | 35.2 (34.4, 36.0) | 141 (140, 143) | 102 (100, 103) | 777 (767, 788) | 0.67 (0.66, 0.68) | 1.97 (1.93, 2.01) | 7.88 (7.83, 7.94) | 5.68 (5.61, 5.75) | 43.4 (43.0, 43.9) |
| 30 | 38.8% (35.8%, 41.8%) | 23.6 (23.2, 24.1) | 17.6 (17.1, 18.2) | 54.7 (53.0, 56.4) | 194 (189, 198) | 141 (138, 145) | 1088 (1060, 1117) | 0.75 (0.73, 0.76) | 2.32 (2.26, 2.37) | 8.21 (8.14, 8.27) | 5.98 (5.91, 6.06) | 46.1 (45.5, 46.6) |
| Lifetime | 52.7% (45.5%, 59.9%) | 27.4 (26.0, 28.8) | 22.1 (20.4, 23.8) | 72.7 (66.6, 78.8) | 232 (217, 246) | 170 (159, 182) | 1310 (1221, 1398) | 0.81 (0.78, 0.83) | 2.65 (2.55, 2.76) | 8.46 (8.34, 8.57) | 6.22 (6.10, 6.34) | 47.8 (46.8, 48.8) |
| **With CVD History: Age 60-70 years** | | | | | | | | | | | | |
| 10 | 21.4% (20.9%, 22.0%) | 9.3 (9.2, 9.3) | 7.9 (7.8, 8.0) | 23.0 (22.6, 23.5) | 82 (82, 83) | 60 (60, 61) | 483 (478, 488) | 0.85 (0.84, 0.87) | 2.49 (2.44, 2.54) | 8.90 (8.84, 8.95) | 6.51 (6.44, 6.59) | 52.1 (51.6, 52.6) |
| 20 | 37.8% (36.3%, 39.3%) | 16.0 (15.8, 16.2) | 15.5 (15.2, 15.9) | 48.4 (47.2, 49.5) | 150 (148, 152) | 111 (109, 113) | 905 (890, 920) | 0.97 (0.95, 0.98) | 3.02 (2.95, 3.09) | 9.36 (9.30, 9.42) | 6.95 (6.87, 7.03) | 56.5 (55.9, 57.1) |
| 30 | 47.9% (45.0%, 50.9%) | 19.5 (19.0, 20.0) | 20.2 (19.4, 21.0) | 65.9 (63.4, 68.5) | 188 (182, 194) | 140 (136, 145) | 1146 (1106, 1186) | 1.04 (1.01, 1.06) | 3.38 (3.29, 3.47) | 9.62 (9.55, 9.70) | 7.20 (7.11, 7.29) | 58.8 (58.0, 59.5) |
| Lifetime | 53.5% (49.2%, 57.8%) | 20.8 (19.9, 21.7) | 22.1 (20.7, 23.5) | 74.2 (69.4, 79.0) | 203 (192, 213) | 152 (144, 161) | 1243 (1173, 1313) | 1.06 (1.04, 1.09) | 3.57 (3.45, 3.68) | 9.75 (9.65, 9.86) | 7.32 (7.20, 7.43) | 59.7 (58.7, 60.7) |

CVD: cardiovascular disease; MVE: major vascular events

***Supplementary table 9. Cumulative effects of statin on CVD events, life years and healthcare use over years***

| **Timeframe (years)** | **Impact of standard statin therapy [Treated - Untreated]** | | | | | | | | | | | |
|---|---|---|---|---|---|---|---|---|---|---|---|---|
| | **Difference in clinical outcomes** | | **Difference in total healthcare use per person** | | | | | **Difference in total healthcare use per person-year** | | | | |
| | **% with new MVE avoided** | **Life years gained per person** | **Hospital admissions** | **Hospital days** | **Consultations** | **Tests** | **Prescription items** | **Hospital admissions** | **Hospital days** | **Consultations** | **Tests** | **Prescription items** |
| **Without CVD History: Age 40-60 years** | | | | | | | | | | | | |
| 10 | 1.2%<br>(1.1%, 1.3%) | 0.00<br>(0.00, 0.01) | -0.02<br>(-0.03, -0.02) | -0.13<br>(-0.14, -0.11) | -0.03<br>(-0.08, 0.01) | 0.00<br>(-0.05, 0.05) | -0.77<br>(-1.09, -0.46) | -0.002<br>(-0.003, -0.002) | -0.013<br>(-0.015, -0.011) | -0.005<br>(-0.010, -0.001) | -0.001<br>(-0.007, 0.004) | -0.085<br>(-0.117, -0.053) |
| 20 | 2.9%<br>(2.6%, 3.1%) | 0.03<br>(0.03, 0.04) | -0.06<br>(-0.07, -0.04) | -0.38<br>(-0.45, -0.31) | 0.07<br>(-0.10, 0.24) | 0.10<br>(-0.10, 0.31) | -2.54<br>(-3.93, -1.14) | -0.004<br>(-0.004, -0.003) | -0.022<br>(-0.025, -0.018) | -0.005<br>(-0.014, 0.004) | 0.000<br>(-0.011, 0.011) | -0.163<br>(-0.237, -0.089) |
| 30 | 4.9%<br>(4.4%, 5.3%) | 0.13<br>(0.09, 0.16) | -0.06<br>(-0.09, -0.03) | -0.65<br>(-0.83, -0.48) | 0.66<br>(0.27, 1.05) | 0.55<br>(0.13, 0.96) | -2.81<br>(-6.06, 0.45) | -0.004<br>(-0.005, -0.003) | -0.029<br>(-0.036, -0.023) | -0.002<br>(-0.015, 0.011) | 0.004<br>(-0.012, 0.019) | -0.201<br>(-0.319, -0.083) |
| Lifetime | 7.5%<br>(6.6%, 8.5%) | 0.64<br>(0.46, 0.81) | 0.33<br>(0.17, 0.48) | 0.44<br>(-0.21, 1.10) | 4.93<br>(3.48, 6.38) | 3.70<br>(2.57, 4.82) | 15.35<br>(6.19, 24.50) | 0.000<br>(-0.002, 0.002)* | -0.016<br>(-0.030, -0.002) | 0.030<br>(0.010, 0.050) | 0.033<br>(0.011, 0.055) | -0.002<br>(-0.196, 0.191) |
| **Without CVD History: Age 60-70 years** | | | | | | | | | | | | |
| 10 | 2.5%<br>(2.2%, 2.7%) | 0.01<br>(0.01, 0.02) | -0.05<br>(-0.05, -0.04) | -0.31<br>(-0.36, -0.27) | -0.06<br>(-0.13, 0.01) | -0.01<br>(-0.09, 0.06) | -1.71<br>(-2.21, -1.20) | -0.005<br>(-0.006, -0.005) | -0.034<br>(-0.039, -0.029) | -0.016<br>(-0.023, -0.009) | -0.007<br>(-0.015, 0.001) | -0.214<br>(-0.267, -0.160) |
| 20 | 5.4%<br>(4.9%, 5.9%) | 0.10<br>(0.08, 0.12) | -0.08<br>(-0.10, -0.05) | -0.79<br>(-0.96, -0.62) | 0.35<br>(0.11, 0.59) | 0.30<br>(0.04, 0.55) | -3.69<br>(-5.65, -1.72) | -0.008<br>(-0.009, -0.006) | -0.053<br>(-0.063, -0.043) | -0.017<br>(-0.031, -0.004) | -0.008<br>(-0.023, 0.008) | -0.362<br>(-0.475, -0.249) |
| 30 | 7.7%<br>(6.9%, 8.5%) | 0.31<br>(0.25, 0.38) | 0.04<br>(-0.03, 0.10) | -0.85<br>(-1.21, -0.48) | 1.96<br>(1.35, 2.57) | 1.46<br>(0.93, 1.99) | 1.00<br>(-3.55, 5.55) | -0.007<br>(-0.009, -0.005) | -0.062<br>(-0.077, -0.047) | -0.009<br>(-0.027, 0.009) | 0.001<br>(-0.019, 0.021) | -0.363<br>(-0.533, -0.193) |
| Lifetime | 8.9%<br>(7.9%, 10.0%) | 0.71<br>(0.57, 0.85) | 0.40<br>(0.26, 0.55) | 0.29<br>(-0.35, 0.93) | 5.58<br>(4.29, 6.86) | 4.12<br>(3.12, 5.12) | 18.61<br>(10.29, 26.94) | -0.003<br>(-0.006, -0.001) | -0.047<br>(-0.067, -0.028) | 0.019<br>(-0.004, 0.041) | 0.027<br>(0.002, 0.051) | -0.157<br>(-0.380, 0.066) |
| **With CVD History: Age 40-60 years** | | | | | | | | | | | | |
| 10 | 4.8%<br>(4.4%, 5.2%) | 0.04<br>(0.03, 0.05) | -0.09<br>(-0.10, -0.07) | -0.59<br>(-0.67, -0.51) | -0.05<br>(-0.19, 0.09) | 0.01<br>(-0.11, 0.14) | -2.92<br>(-3.99, -1.86) | -0.011<br>(-0.013, -0.010) | -0.068<br>(-0.076, -0.059) | -0.035<br>(-0.046, -0.024) | -0.020<br>(-0.031, -0.009) | -0.459<br>(-0.549, -0.369) |
| 20 | 8.0%<br>(7.3%, 8.8%) | 0.19<br>(0.15, 0.23) | -0.09<br>(-0.14, -0.04) | -1.08<br>(-1.33, -0.82) | 1.01<br>(0.53, 1.48) | 0.80<br>(0.38, 1.22) | -2.62<br>(-6.37, 1.14) | -0.012<br>(-0.014, -0.010) | -0.081<br>(-0.094, -0.067) | -0.029<br>(-0.046, -0.012) | -0.017<br>(-0.035, 0.002) | -0.610<br>(-0.772, -0.448) |
| 30 | 9.8%<br>(8.8%, 10.8%) | 0.47<br>(0.36, 0.58) | 0.09<br>(-0.03, 0.21) | -0.87<br>(-1.39, -0.35) | 3.51<br>(2.41, 4.62) | 2.69<br>(1.79, 3.58) | 7.78<br>(-0.49, 16.04) | -0.011<br>(-0.014, -0.008) | -0.081<br>(-0.099, -0.063) | -0.014<br>(-0.036, 0.008) | -0.005<br>(-0.029, 0.019) | -0.575<br>(-0.800, -0.350) |
| Lifetime | 10.2%<br>(8.8%, 11.6%) | 0.94<br>(0.70, 1.17) | 0.60<br>(0.33, 0.88) | 0.89<br>(-0.14, 1.92) | 8.35<br>(5.94, 10.76) | 6.39<br>(4.51, 8.27) | 33.55<br>(17.78, 49.32) | -0.005<br>(-0.009, -0.001) | -0.056<br>(-0.080, -0.032) | 0.015<br>(-0.012, 0.042) | 0.020<br>(-0.009, 0.049) | -0.396<br>(-0.677, -0.115) |
| **With CVD History: Age 60-70 years** | | | | | | | | | | | | |
| 10 | 7.1%<br>(6.5%, 7.7%) | 0.06<br>(0.05, 0.07) | -0.12<br>(-0.14, -0.10) | -0.93<br>(-1.08, -0.78) | -0.04<br>(-0.20, 0.13) | 0.07<br>(-0.08, 0.22) | -4.12<br>(-5.41, -2.84) | -0.018<br>(-0.021, -0.016) | -0.117<br>(-0.133, -0.100) | -0.063<br>(-0.078, -0.048) | -0.036<br>(-0.051, -0.021) | -0.788<br>(-0.911, -0.665) |
| 20 | 10.6%<br>(9.6%, 11.6%) | 0.28<br>(0.23, 0.33) | -0.05<br>(-0.12, 0.03) | -1.35<br>(-1.76, -0.94) | 1.64<br>(1.03, 2.25) | 1.37<br>(0.86, 1.88) | -1.14<br>(-5.95, 3.67) | -0.019<br>(-0.023, -0.016) | -0.135<br>(-0.159, -0.110) | -0.059<br>(-0.082, -0.036) | -0.035<br>(-0.059, -0.010) | -1.034<br>(-1.254, -0.814) |
| 30 | 11.6%<br>(10.4%, 12.8%) | 0.56<br>(0.44, 0.68) | 0.24<br>(0.08, 0.39) | -0.75<br>(-1.47, -0.03) | 4.48<br>(3.19, 5.77) | 3.55<br>(2.54, 4.56) | 12.35<br>(2.67, 22.03) | -0.017<br>(-0.021, -0.013) | -0.132<br>(-0.162, -0.101) | -0.045<br>(-0.073, -0.017) | -0.024<br>(-0.053, 0.005) | -1.023<br>(-1.313, -0.734) |
| Lifetime | 11.5%<br>(10.2%, 12.8%) | 0.80<br>(0.62, 0.97) | 0.54<br>(0.30, 0.78) | 0.34<br>(-0.66, 1.34) | 7.16<br>(5.20, 9.12) | 5.62<br>(4.10, 7.14) | 27.46<br>(13.72, 41.20) | -0.014<br>(-0.019, -0.009) | -0.115<br>(-0.150, -0.081) | -0.027<br>(-0.058, 0.003) | -0.009<br>(-0.041, 0.023) | -0.927<br>(-1.252, -0.602) |

CVD: cardiovascular disease; MVE: major vascular events

*Results over time: 0.0000002 (-0.002, 0.002) over lifetime (year 70), -0.0000007 (-0.002, 0.002) over year 69, and -0.002 (-0.004, -0.0001) over year 47 (last year with statistically significant rate reduction since entry).

*Supplementary table 10. Time to neutral net effects of statin on healthcare use per person*

| Subgroup | | Time since statin initiation to neutral net effect, mean (95%CIs) years | | | | |
|---|---|---|---|---|---|---|
| CVD history | Age (years) | Hospital admissions | Hospital days | Consultations | Tests | Prescription items |
| No | 40-60 | 38 (33, 44) | 51 (43, 62) | 17 (9, 25) | 10 (4, 27) | 38 (28, 48) |
| | 60-70 | 29 (25, 32) | 43 (36, 49) | 14 (10, 18) | 12 (6, 19) | 29 (24, 36) |
| Yes | 40-60 | 27 (23, 32) | 41 (34, 52) | 12 (9, 15) | 10 (7, 15) | 24 (19, 31) |
| | 60-70 | 22 (19, 27) | 40 (30, 47) | 11 (9, 14) | 9 (7, 12) | 22 (17, 28) |

CVD: cardiovascular disease;

*Supplementary figure 6. Effects of statin treatment on incident cancer and diabetes (among people without such condition at statin initiation) over time*

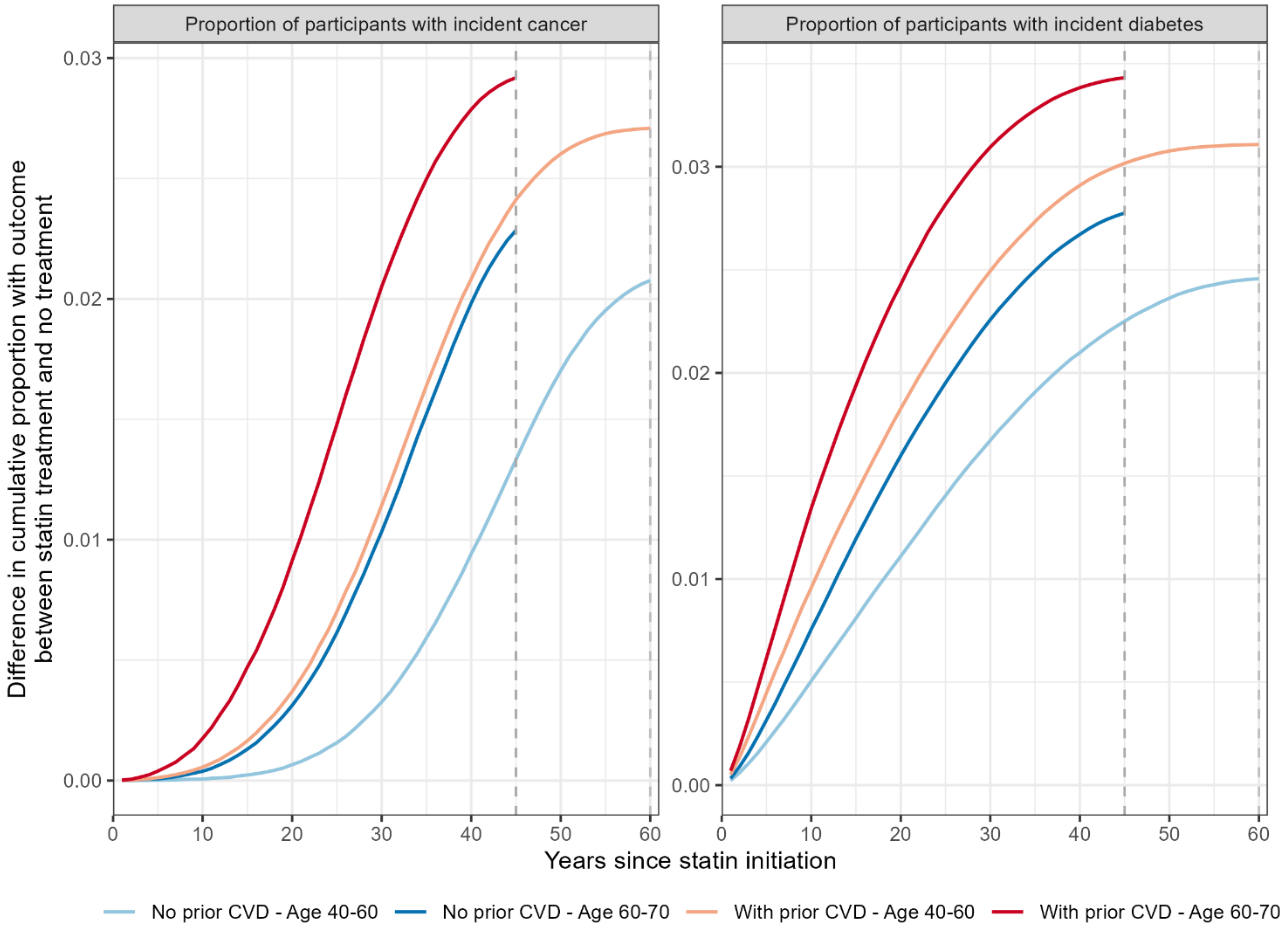

Lines represents the mean estimates. Assuming all participants died at age 110 years, the summary ends in year 60 for people aged 40-60 and in year 45 for people aged 60-70.

### Supplementary references

1. Law MR, Wald NJ, Rudnicka AR. Quantifying effect of statins on low density lipoprotein cholesterol, ischaemic heart disease, and stroke: systematic review and meta-analysis. *BMJ* 2003; **326**(7404): 1423.
2. Cholesterol Treatment Trialists C, Mihaylova B, Emberson J, et al. The effects of lowering LDL cholesterol with statin therapy in people at low risk of vascular disease: meta-analysis of individual data from 27 randomised trials. *Lancet* 2012; **380**(9841): 581-90.
3. Sattar N, Preiss D, Murray HM, et al. Statins and risk of incident diabetes: a collaborative meta-analysis of randomised statin trials. *Lancet* 2010; **375**(9716): 735-42.
4. Preiss D, Seshasai SR, Welsh P, et al. Risk of incident diabetes with intensive-dose compared with moderate-dose statin therapy: a meta-analysis. *JAMA* 2011; **305**(24): 2556-64.
5. Law M, Rudnicka AR. Statin safety: a systematic review. *Am J Cardiol* 2006; **97**(8A): 52C-60C.
